# A next-generation electronic frailty index leveraging deep learning on unstructured health records extends risk prediction across the full frailty spectrum

**DOI:** 10.64898/2026.06.23.26356323

**Authors:** Esha Khan, Silvia Ottaviani, Juho Kaijansinkko, Markus J Haapanen, Anna Tirkkonen, Jonathan K L Mak, Hanna Pajulammi, Mikaela B von Bonsdorff, Jake Lin, Juulia Jylhävä

**Affiliations:** Faculty of Medicine and Health Technology, Tampere University, Tampere, Finland; Geriatric Clinic, Department of Internal Medicine and Medical Specialties, University of Genoa, Genoa, Italy; Research Unit of Clinical Medicine, Faculty of Medicine, University of Oulu, Oulu, Finland; Folkhälsan Research Center, Helsinki, Finland; Centre for Health Services Research, Faculty of Medicine, The University of Queensland, Princess Alexandra Hospital, Woolloongabba, Queensland, Australia; Faculty of Sport and Health Sciences and Gerontology Research Center, University of Jyväskylä, Jyväskylä, Finland; Department of Medical Epidemiology and Biostatistics, Karolinska Institutet, Stockholm, Sweden; Department of Pharmacology and Pharmacy, Li Ka Shing Faculty of Medicine, The University of Hong Kong, Hong Kong, China; Department of Geriatric Medicine, Central Finland Hospital Nova, Wellbeing Services County of Central Finland, Finland; Faculty of Medicine, University of Helsinki, Helsinki, Finland; Research Unit of Population Health, Faculty of Medicine, University of Oulu, Oulu, Finland

## Abstract

**Background:** Existing electronic frailty indices (eFI) are typically based on structured data and designed for older adults. We developed an eFI that integrates structured and unstructured electronic health records (EHRs) across adulthood and assessed its longitudinal trajectories and associations with adverse outcomes.

**Methods:** We used longitudinal EHR data from 193629 individuals aged 35–103 in the Wellbeing Services County of Central Finland (2010–2023) and constructed a 53-item eFI including diagnosis codes, laboratory tests and items extracted from free-text clinical notes using deep-learning-based natural language processing. Associations with all-cause mortality, severe infections, fractures, and healthcare utilization were assessed using Cox and count models. Predictive performance was compared with Hospital Frailty Risk Score (HFRS) and Charlson Comorbidity Index (CCI).

**Findings:** eFI trajectories accelerated notably from age 65 onwards. Using the eFI as a categorical variable, severe frailty was associated with higher risks of mortality (hazard ratio [HR] 7·31, 95% confidence interval [CI] 6·83–7·83), severe infections (HR 9·22, 95%CI 8·52– 9·98), fractures (HR 2·75, 95%CI 2·52–3·01) and increased healthcare utilization (odds ratio [OR] 3·15, 95%CI 2·96–3·35) compared with non-frail. The risks were relatively greater in younger age groups and persisted when using the continuous eFI restricted to non-frail individuals. Across all outcomes, the eFI showed greater model discrimination than HFRS and CCI.

**Interpretation:** An eFI using structured and unstructured EHR data improves risk stratification even in younger adults and at very low levels of frailty.

**Funding:** Research Council of Finland, Instrumentarium Science Foundation, Sigrid Jusélius Foundation and Samfundet Folkhälsan.

## Introduction

Frailty is a clinical state of increased vulnerability resulting from a decline in physiological reserves and a reduced capacity to maintain homeostasis in response to stress.^1,2^ It has emerged as a key determinant of various adverse outcomes, such as mortality, healthcare utilization, and admission to long-term care,^3,4^ yet its routine assessment in clinical practice remains limited. Many existing frailty models, such as the frailty index (FI)^2^ and frailty phenotype are comprehensive in research settings but they are difficult to implement at scale in real-world clinical environments.^5^ The FI, which conceptualizes frailty as the cumulative burden of health deficits across physiological systems can nevertheless be implemented within electronic health records (EHR) as an electronic FI (eFI).^6^

eFI models have been developed in several countries and demonstrated promising predictive performance and clinical utility.^7^ However, most of these studies have been restricted to older populations, relied only on structured and/or cross-sectional data, and specific care settings such as primary care, hospitalized patients or surgical cohorts.^8–10^ As a result, little is known about how the eFI performs in younger individuals, despite evidence that frailty is already present and clinically meaningful in midlife,^11,12^ or about how eFI trajectories evolve over time in clinical populations. Moreover, eFIs have been successfully implemented only few countries, such as the UK,^13^ and its wider adoption elsewhere has been limited in part by differences in underlying EHR coding systems. The UK eFI relies on Read Codes, which include detailed representations of clinical manifestations, symptoms, functional problems, and other health issues commonly documented in primary care. However, Read Codes are not used in other countries, where diagnoses are typically recorded using International Classification of Diseases (ICD) -based coding systems, limiting the direct transferability of the UK eFI to other countries. As a result, several health deficits included in the UK eFI cannot be directly replicated elsewhere. Moreover, relying solely on structured ICD-coded data may lead to many clinically relevant features of frailty, such as functional limitations, sensory functions, and psychosocial wellbeing, being incompletely captured, since they are typically documented only in free-text clinical notes.

Because of these limitations, no universal eFI model applicable across all healthcare settings has yet been established. Artificial intelligence (AI)-driven methods for extracting information from free-text clinical notes, however, now allow incorporation of unstructured EHR data at scale, offering a more comprehensive assessment of health deficits than structured data alone.^14,15^ These methods can also capture earlier indications of health decline not yet recorded in structured data.

In this study, we developed a next-generation eFI within a large Finnish EHR dataset covering primary, secondary and tertiary care, aiming to: (1) construct an eFI integrating structured clinical data and detailed, frailty-related information derived from unstructured health records using deep-learning AI; (2) describe longitudinal patterns of eFI across midlife and older adulthood, and (3) evaluate associations between the eFI and adverse health outcomes, including mortality, severe infections, fractures, and healthcare utilization; (4) compare its predictive performance with established measures, such as the Hospital Frailty Risk Score (HFRS) and Charlson Comorbidity Index (CCI). We also extended the analysis by restricting it to individuals classified as non-frail, focusing on the lower end of the eFI spectrum to investigate health risks associated with low levels of frailty, an area that has remained unexplored in previous research.

### Research in context

#### Evidence before this study

We searched MEDLINE/PubMed for studies published up to Feb 23, 2026, without specifying an inception date, using the terms (“electronic frailty index” OR “frailty index”) AND (“infection” OR “fracture” OR “healthcare utilization” OR “mortality”). One review identified multiple electronic frailty index (eFI) approaches across diverse settings, yet most were applied in selected environments with relatively limited sample sizes, underscoring both the broad utility of the eFIs and the heterogeneity in how they have been constructed and applied. When compared to in-person assessment, another review found that the accuracy of electronic frailty measures in primary care varied considerably. Large cohort studies among older adults have shown that higher frailty levels are associated with an increased risk of adverse outcomes, mainly mortality and hospitalization. However, most studies rely on structured data and cross-sectional assessments, typically do not capture information from free-text clinical records, and are largely restricted to older populations with higher levels of frailty. Consequently, little is known about whether variation within the non-frail range of frailty reflects meaningful differences in health risk, and whether even very low levels of frailty carry prognostic significance.

#### Added value of this study

Combining electronic health record (EHR) data from primary, secondary, and tertiary care, we developed an eFI that integrates structured clinical data on diagnoses and laboratory tests and information derived from free-text clinical records. The eFI comprised 53 items, including 10 deficits identified from the free texts (mobility limitations, physical and cognitive functioning, sensory functions, falls, incontinence, and psychosocial wellbeing) using an AI-driven deep-learning approach, and was applied across the adult age spectrum and modelled for longitudinal trajectories. We evaluated its association with mortality, severe infections, fractures, and healthcare utilization, and compared its predictive performance with established risk scores. This approach extends previous work by incorporating unstructured data recorded by all healthcare professionals, including younger adults (aged 35+), assessing a range of clinically relevant outcomes, and specifically examining prognostic utility within the non-frail subgroup. Moreover, our data represent a real-world, unselected population, in contrast to many cohort, survey, or clinically defined EHR datasets, which often rely on selected samples, specific patient groups, or are shaped by insurance-based access. In the Nordic welfare model, universal entitlement to public healthcare ensures that inclusion is not conditioned by socioeconomic status, thereby reducing selection bias.

#### Implications of all the available evidence

Our findings indicate that an eFI constructed from routinely collected structured and unstructured EHR data serves as a risk indicator across adulthood, with higher frailty conferring relatively greater risk for adverse health outcomes among younger adults. The consistent dose–response associations with the adverse outcomes across the age range and frailty spectrum, including at very low levels of frailty, indicate that it captures a clinically meaningful gradient of vulnerability. This supports the use of the eFI to more accurately identify at-risk individuals across the frailty continuum and adult age range, and to guide more personalized care, including earlier prevention of adverse events and the better alignment of healthcare intensity with patients’ underlying reserves.

## Methods

### Study design and population

We conducted a population-based retrospective study using real-world EHR data from the Wellbeing Services County of Central Finland, the eleventh largest of Finland’s 21 wellbeing services counties, serving approximately 270,000 inhabitants across urban and rural areas.^16^ The dataset included all inpatient and outpatient EHRs from public primary, secondary, tertiary, long-term, and home-care services recorded by all healthcare professionals between 1 January 2010 and 31 December 2023.

Available information included ICD-10 -coded diagnoses, routine laboratory results, procedures recorded using Nordic Medico-Statistical Committee (NOMESCO) procedure codes, prescribed medications, and free-text notes. The free-text notes covered clinical notes (e.g., progress notes, admission and discharge summaries, operative and consultation reports), physical examination findings, nursing documentation, assessments of social circumstances, physiotherapy and rehabilitation notes, psychiatric evaluations, radiology reports, care plans, and patient or family communications, including telephone encounters. In total, the study population comprised 201,571 individuals born between 1920 and 1975, whose baseline ages (i.e., age at first EHR entry) ranged from 35 to 103 years. The eFI was constructed for all available individuals present in the dataset. Entry year was defined as the first calendar year in which a person appeared in the data. Individuals were followed up from their entry year until death or the last available EHR. A flowchart of the eFI construction is presented in the Supplementary Figure 1 (appendix p 18).

Informed consent and review and/or approval by an ethics committee were not required for this study as according to the Finnish legislation (Act on the Secondary Use of Health and Social Data 552/2019 by the Ministry of Social Affairs and Health), the patients included in the study sample were not contacted, and the study did not affect the treatment of the patients. The Human Sciences Ethics Committee of the University of Jyväskylä has certified these conditions pertinent to our study and stated that an ethical review is not required. The study was conducted in accordance with the Helsinki Declaration and in accordance with all relevant guidelines and regulations.

### eFI deficits

The eFI was constructed according to the deficit accumulation model of frailty, following established criteria for FI construction.^17^ The eFI items (deficits) were selected based on clinical relevance and coded as binary variables (0=absent, 1=present), and weighted equally. The eFI comprised 53 items as shown in Supplementary Table 1 (appendix p 3), including ICD-10-coded diagnoses (N = 34), laboratory tests (N = 9), and free-text-derived deficits identified using deep-learning AI-driven natural language processing (NLP) (N = 10). The free-text–derived deficits included falls, incontinence, loneliness, mobility limitations, hearing impairment, visual impairment, age-related neurocognitive problems and the need for assistance with bathing, dressing, and eating or preparing meals (Supplementary Table 1, appendix p 3). Details of the eFI item ascertainment, and descriptions of HFRS and CCI calculation are provided in Supplementary Methods (appendixp 1).

### Outcomes

We selected all-cause mortality, severe infections requiring hospital treatment, and healthcare utilization as outcomes to capture diverse, clinically relevant domains of vulnerability. Severe infections represent impaired immune competence, consistent with evidence linking frailty to chronic systemic inflammation;^18^ fractures represent a well-established consequence of physiological vulnerability and impaired musculoskeletal integrity; and mortality and healthcare utilization reflect diminished overall resilience and reduced capacity to withstand physiological stressors.

Follow-up for all-cause mortality began on January 1^st^ of the year following the baseline eFI period (e.g., for a person having the baseline eFI calculated in 2010–2012, follow-up started January 1^st^, 2013).

Severe infections and fractures were identified using predefined ICD-10 codes as detailed in the Supplementary Table 2 (appendix p 5). Individuals were followed for two years after the baseline eFI period to assess incident severe infections and fractures. Severe infections and fractures were analysed as separate outcomes, both as a time-to-event outcome (any severe infection or fracture within two years after baseline eFI assessment) and count outcome (number of severe infections or fracture episodes in the same period). Subjects without events were censored at two years of follow-up or death, whichever occurred first.

Healthcare utilization was assessed during the first year following the baseline eFI assessment, defined as the total number of visits to primary and secondary care, a timeframe commonly used in health-services research. Subjects who did not use healthcare during this period were assigned a value of zero.

Supplementary Figure 2 (appendix p 19) illustrates the outline of follow-up time, and Supplementary Figure 3 (appendix p 20) reports the sample sizes for each outcome analysis.

### Statistical Analysis

Baseline characteristics were summarized using medians and interquartile ranges (IQRs) or means and standard deviations (SDs) for continuous variables, as appropriate, and counts with percentages for categorical variables. Correlations across individual eFI items were assessed using Pearson correlation coefficients to determine whether each deficit contributed unique information to the eFI. The correlations between baseline eFI and HFRS and CCI was assessed using Spearman’s rank correlation coefficient.

To examine changes in frailty over time, annual eFI values were calculated for each individual for every calendar year. Longitudinal trajectories of the eFI were visualized using spaghetti plots using a random sample of 50000 participants per sex, with locally estimated scatterplot smoothing curves. The eFI trajectories by sex were estimated using generalized additive mixed models (GAMMs) with a Tweedie distribution and a log link. Age was modelled as a penalized spline with sex-specific smooth terms with a random intercept and slope included to account for repeated measurements.

Cox proportional hazards models were fitted to examine associations between baseline eFI and time-to-event outcomes, i.e., mortality, severe infections, and fractures. The analyses were performed in the full sample and additionally stratified by sex and age group (35–49, 50–64, 65–79, ≥80). To examine the risks explicitly at the lower end of the eFI spectrum, we also performed the analyses only among individuals classified as non-frail.

A base model including baseline age and sex was fitted first; baseline eFI, CCI, and HFRS were then added individually to this model to assess and compare prognostic performance. Model discrimination was evaluated using Harrell’s concordance index (C-index), and improvements over the base model were calculated for the eFI, CCI, and HFRS. Proportional hazards’ assumptions were assessed and confirmed using Schoenfeld residuals. The generalizability of the Cox models was evaluated using a 70:30 train–test split (hold-out validation) with predictive performance (C-index) assessed on the held-out test set. Methods for count models on severe infections, fracture episodes, and healthcare utilization are provided in Supplementary Methods (appendix p 1). All statistical analyses were conducted using R version 4.5.2. Statistical significance was set at a two-sided α of 0·05.

## Results

### Sample characteristics

As shown in Table 1 and Supplementary Figure 1, 193629 subjects were included, where 34727 subjects had only one eFI assessment across the years, of whom 51·6% were female. The mean age at baseline was 62·0 years (SD 13·6). The median eFI was 2·3 (IQR 7·7) and was similar in females and men. The proportion of females was higher in the oldest age group (≥80 years: 14·7% vs 9·6%).

**Table 1.** Baseline characteristics of the population. Continuous variables are summarized as mean and standard deviation (SD) and median and quartiles (Q) as appropriate. Categorical variables are presented as n (%). For binary variables, Yes and No indicate presence and absence of the event in the defined observation period. P-values indicate differences between females and males using the Wilcoxon rank-sum test for continuous variables and Pearson’s chi–squared (χ^2^) test for categorical variables.

|  | **Overall**<br>**N = 193629** | **Females**<br>**N = 99837** | **Males**<br>**N = 93792** | **p-value** |
| --- | --- | --- | --- | --- |
| **eFI** |  |  |  | <0.0001 |
| Mean ± SD | 4.9 ± 6.2 | 5.0 ± 6.2 | 4.9 ± 6.1 | |
| Median (Q1, Q3) | 2.3 (0.0, 7.7) | 2.3 (0.0, 7.7) | 2.3 (0.0, 7.7) |  |
| eFI categories, N (%) |  |  |  | <0.0001 |
| Non-frail ($\leq 8$) | 149090 (77.0) | 77064 (77.2) | 72026 (76.8) | |
| Mild Frailty (>8–15) | 27808 (14.4) | 13973 (14.0) | 13835 (14.8) |  |
| Moderate Frailty (>15–21) | 11471 (5.9) | 5910 (5.9) | 5561 (5.9) |  |
| Severe Frailty (>21) | 5260 (2.7) | 2890 (2.9) | 2370 (2.5) |  |
| **Age** |  |  |  | <0.0001 |
| Mean ± SD | 62.0 ± 13.6 | 62.8 ± 14.1 | 61.2 ± 13.0 | |
| Median (Q1, Q3) | 62.0 (51.0, 72.0) | 62.0 (52.0, 73.0) | 61.0 (51.0, 70.0) |  |
| Age group, N (%) |  |  |  | <0.0001 |
| 35–49 | 40858 (21.1) | 20437 (20.5) | 20421 (21.8) |  |
| 50–64 | 70807 (36.6) | 35058 (35.1) | 35749 (38.1) |  |
| 65–79 | 58316 (30.1) | 29699 (29.7) | 28617 (30.5) |  |
| $\geq 80$ | 23648 (12.2) | 14643 (14.7) | 9005 (9.6) | |
| **HFRS** |  |  |  | <0.0001 |
| Mean ± SD | 0.8 ± 1.9 | 0.9 ± 2.0 | 0.8 ± 1.8 | |
| Median (Q1, Q3) | 0.0 (0.0, 0.9) | 0.0 (0.0, 1.1) | 0.0 (0.0, 0.8) |  |
| HFRS categories, N (%) |  |  |  | <0.0001 |
| Low risk ($< 5$) | 186314 (96.2) | 95813 (96.0) | 90501 (96.5) | |
| Intermediate risk (5–15) | 6923 (3.6) | 3805 (3.8) | 3118 (3.3) |  |
| High risk ($> 15$) | 392 (0.2) | 219 (0.2) | 173 (0.2) | |
| **CCI** |  |  |  | <0.0001 |
| Mean ± SD | 0.3 ± 0.9 | 0.3 ± 0.8 | 0.4 ± 0.9 | |
| Median (Q1, Q3) | 0.0 (0.0, 0.0) | 0.0 (0.0, 0.0) | 0.0 (0.0, 0.0) |  |
| CCI categories, N (%) |  |  |  | <0.0001 |
| No comorbidity (=0) | 157670 (81.4) | 81680 (81.8) | 75,990 (81.0) |  |
| Mild comorbidity (=1) | 18246 (9.4) | 9864 (9.9) | 8382 (8.9) |  |
| Moderate comorbidity (=2) | 12928 (6.7) | 6354 (6.4) | 6574 (7.0) |  |
| Severe comorbidity ($\geq 3$) | 4785 (2.5) | 1939 (1.9) | 2846 (3.0) | |
| **Death, N (%)** |  |  |  | <0.0001 |
| No | 155023 (80.1) | 81074 (81.2) | 73949 (78.8) |  |
| Yes | 38606 (19.9) | 18763 (18.8) | 19843 (21.2) |  |
| **Severe infections, N (%)** |  |  |  | 0.12 |
| No | 187136 (96.6) | 96552 (96.7) | 90584 (96.6) |  |
| Yes | 6493 (3.4) | 3285 (3.3) | 3208 (3.4) |  |
| **Fractures, N (%)** |  |  |  | <0.0001 |
| No | 183754 (94.9) | 93951 (94.1) | 89803 (95.7) |  |
| Yes | 9875 (5.1) | 5886 (5.9) | 3989 (4.3) |  |
| **Healthcare utilization, N (%)** |  |  |  | <0.0001 |
| No | 100221 (51.8) | 48430 (48.5) | 51791 (55.2) |  |
| Yes | 93408 (48.2) | 51407 (51.5) | 42001 (44.8) |  |

CCI, Charlson Comorbidity Index; eFI, electronic frailty index; HFRS, Hospital Frailty Risk Score.

Three cut points (8, 15, and 21) were identified for the eFI using the data-driven approach on observed mortality risk, defining the frailty categories as follows: non-frail (≤8), mild frailty (>8–15), moderate frailty (>15–21), and severe frailty (>21). Overall, 149090 (77·0%) of subjects were classified as non-frail by the eFI categories, 27808 (14·4%) as mildly frail, 11471 (5·9%) as moderately frail, and 5260 (2·7%) as severely frail. Severe frailty was most common in the two oldest age groups (65–79 years: 35·1% and ≥80 years: 51·3%), but it was also present in younger age groups (35–49 years: 2·0% and 50–64 years: 11·6%) (Supplementary Table 3, appendix p 6). Age, HFRS, and CCI increased progressively across the eFI categories (Supplementary Table 3, appendix p 6). The eFI had a moderate correlation with both HFRS (ρ = 0·47) and CCI (ρ = 0·49).

The NLP–NER models showed good overall performance in identifying deficit items from unstructured free texts, with F1 scores ranging from 0·74 to 0·92 across items (Supplementary Table 4, appendix p 7). Correlations among the eFI items were generally weak to moderate for laboratory items (maximum ρ =0·56 between fasting plasma glucose and glycated haemoglobin) and weaker for free-text and diagnosis-based items (Supplementary Table 5, appendix p 8), indicating that all items contributed unique information to the eFI.

### eFI trajectories

Frailty increased with age in both sexes, following a non-linear pattern with a marked acceleration in the rate of increase around age 65 (Figure 1). The trajectories were largely similar between females and males across most of the age range. The GAMMs confirmed a significant non-linear association between age and eFI, with sex-specific smooth terms indicating modest, but statistically significant differences in the eFI trajectories between females and males (Supplementary Table 6, appendix p 9).

**Figure 1.**
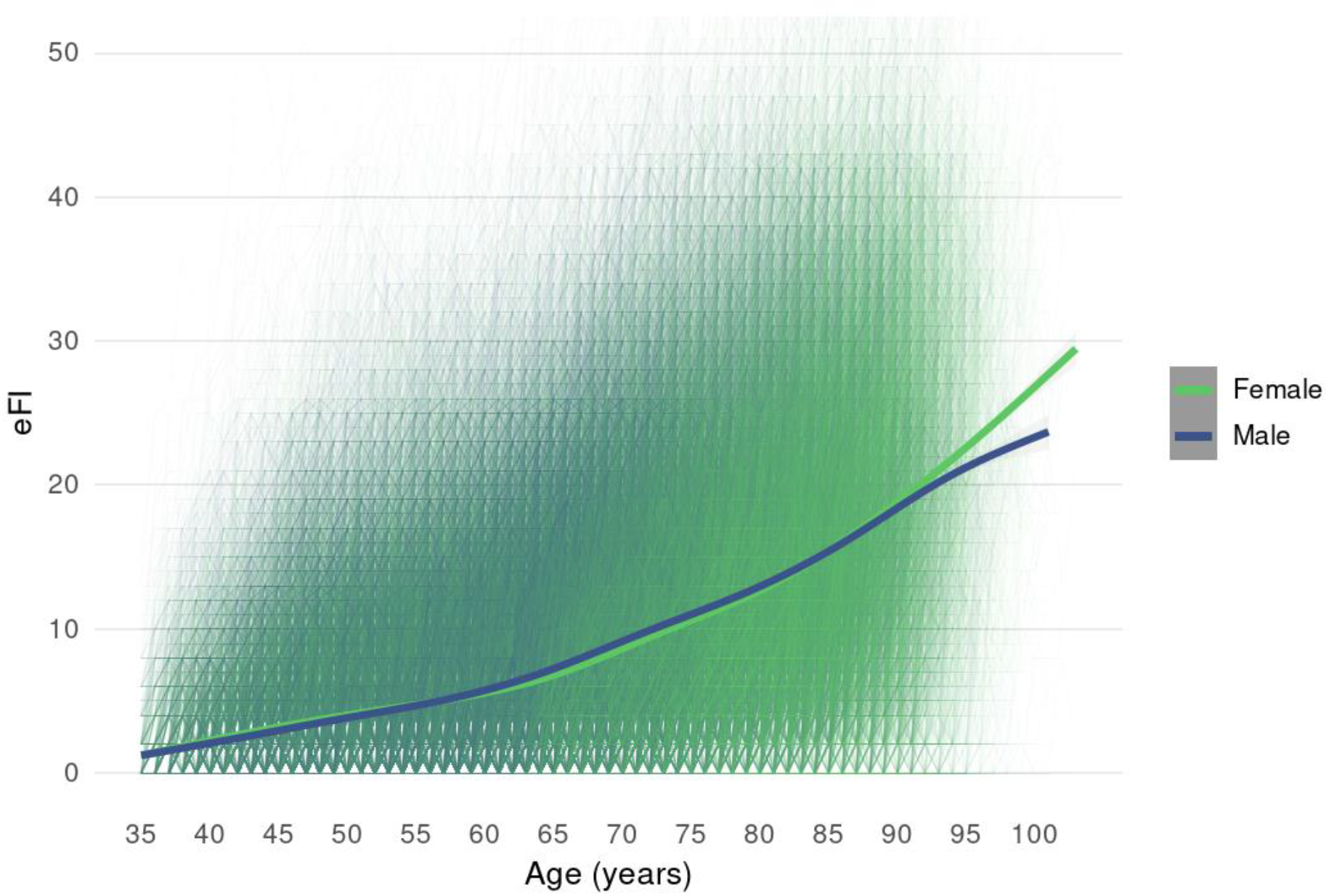
Longitudinal frailty trajectories. Individual-level trajectories are shown as semi-transparent (spaghetti) lines. Sex-specific smoothed trends are shown with the bolded lines for females (green) and males (blue), estimated using generalized additive models with spline functions for age, The 95% confidence intervals around the trend lines are shown in grey.

**Figure 2.**
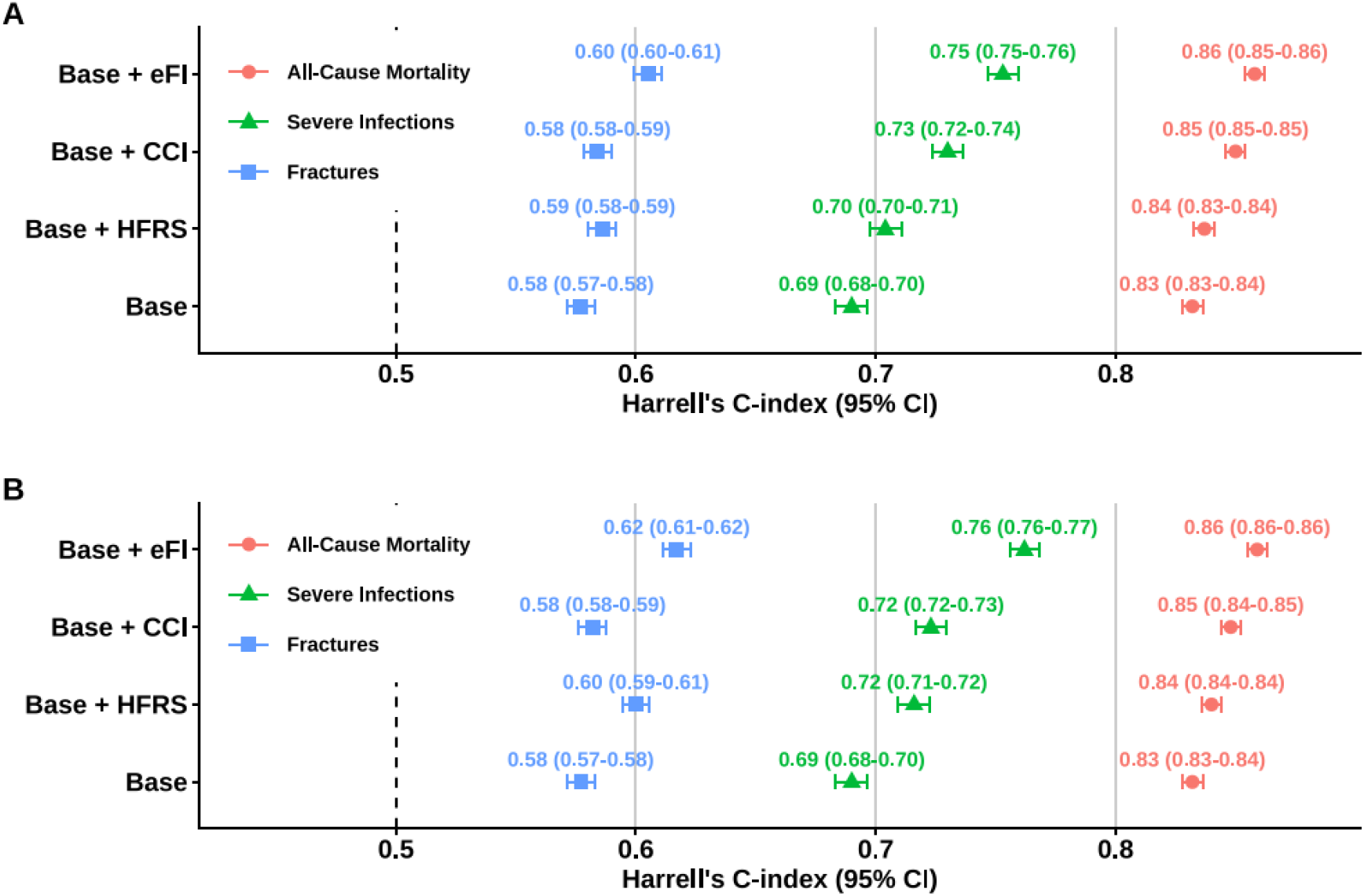
Model discrimination based on concordance (C-index) and its 95%CI for all-cause mortality, severe infections, and fractures. The eFI, CCI, and HFRS were each added individually to the base model containing age and sex. In panel A, eFI, CCI, and HFRS were modelled as categorical variables, whereas in panel B they were modelled as continuous variables. CCI = Charlson Comorbidity Index; CI = Confidence Interval; eFI = electronic Frailty Index; HFRS = Hospital Frailty Risk Score.

### Associations with outcomes

Higher baseline eFI was consistently associated with increased risk of the time-to-event outcomes (all-cause mortality, severe infections, and fractures) in the Kaplan–Meier analyses (Supplementary Figure 4, appendix p 21). In the Cox models, the risk of all-cause mortality increased in a dose-responsive manner across the eFI categories (mild frailty: HR 2·14, 95% CI 2·03–2·26; moderate frailty: HR 3·97, 95% CI 3·74–4·21; severe frailty: HR 7·31, 95% CI 6·83–7·83; continuous eFI: HR 1·09, 95% CI 1·09–1·10) (Table 2). For severe infections, progressively higher risks were observed with increasing risk across the eFI categories (mild frailty: HR 2·48, 95% CI 2·32–2·65; moderate frailty: HR 4·73, 95% CI 4·41–5·08; severe frailty: HR 9·22, 95% CI 8·52–9·98; continuous eFI: HR 1·10, 95% CI 1·10–1·10) (Table 2). For fractures, the risks similarly increased across the eFI categories (mild: HR 1·55, 95% CI 1·47–1·64; moderate: HR 2·11, 95% CI 1·98–2·26; severe: HR 2·75, 95% CI 2·52–3·01; continuous eFI: HR 1·05, 95% CI 1·05–1·05) (Table 2). HFRS and CCI were also associated with all outcomes, across both categorical and continuous definitions (Table 2).

**Table 2.** Associations of eFI, CCI and HFRS with all-cause mortality, severe infections, and fractures using Cox regression and healthcare utilization using hurdle negative binomial models.

|  | **All-cause**<br>**Mortality**<br>**HR (95% CI)**<br>**(N = 124874;**<br>**events = 8923)** | **Severe Infections**<br>**HR (95% CI)**<br>**(N = 142229;**<br>**events = 6493)** | **Fractures**<br>**HR (95% CI)**<br>**(N = 142209;**<br>**events = 9875)** | **Healthcare**<br>**utilization OR**<br>**(95% CI)**<br>**(N = 178163;**<br>**events = 93408)** | **Healthcare**<br>**utilization IRR**<br>**(95% CI)**<br>**(N = 178163)** |
| --- | --- | --- | --- | --- | --- |
| **eFI** |  |  |  |  |  |
| Categorical |  |  |  |  |  |
| Non-frail ($\leq 8$, Ref.) | 1 | 1 | 1 | 1 | 1 |
| Mild frailty ($> 8-15$) | 2.14 (2.03–2.26) | 2.48 (2.32–2.65) | 1.55 (1.47–1.64) | 2.62 (2.55, 2.69) | 1.91 (1.87, 1.96) |
| Moderate frailty ($> 15-21$) | 3.97 (3.74–4.21) | 4.73 (4.41–5.08) | 2.11 (1.98–2.26) | 3.56 (3.41, 3.72) | 2.85 (2.76, 2.94) |
| Severe frailty ($> 21$) | 7.31 (6.83–7.83) | 9.22 (8.52–9.98) | 2.75 (2.52–3.01) | 3.15 (2.96, 3.35) | 3.98 (3.80, 4.16) |
| Continuous per point increase | 1.09 (1.09–1.10) | 1.10 (1.10–1.10) | 1.05 (1.05–1.05) | 1.12 (1.11, 1.12) | 1.08 (1.08, 1.08) |
| **HFRS** |  |  |  |  |  |
| Categorical |  |  |  |  |  |
| Low risk ($< 5$, Ref.) | 1 | 1 | 1 | 1 | 1 |
| Intermediate risk ($5-15$) | 2.67 (2.50–2.85) | 2.81 (2.61–3.02) | 1.88 (1.74–2.02) | 2.21 (2.10, 2.33) | 2.09 (2.00, 2.17) |
| High risk ($> 15$) | 3.91 (3.12–4.89) | 3.72 (2.97–4.65) | 2.69 (2.09–3.46) | 2.66 (2.12, 3.34) | 2.29 (1.94, 2.70) |
| Continuous per point increase | 1.12 (1.12–1.13) | 1.13 (1.12–1.13) | 1.09 (1.08–1.10) | 1.16 (1.15, 1.16) | 1.14 (1.13, 1.14) |
| **CCI** |  |  |  |  |  |
| Categorical |  |  |  |  |  |
| None (0, Ref.) | 1 | 1 | 1 | 1 | 1 |
| Mild (1) | 2.05 (1.94–2.16) | 2.39 (2.25–2.55) | 1.32 (1.25–1.40) | 1.82 (1.76, 1.88) | 1.63 (1.59, 1.68) |
| Moderate (2) | 2.75 (2.59–2.92) | 2.76 (2.57–2.97) | 1.21 (1.12–1.30) | 1.51 (1.45, 1.57) | 2.07 (2.01, 2.14) |
| Severe ($\geq 3$) | 5.18 (4.80–5.60) | 5.01 (4.59–5.47) | 1.77 (1.59–1.98) | 1.53 (1.44, 1.62) | 3.54 (3.36, 3.73) |
| Continuous per point increase | 1.48 (1.46–1.50) | 1.43 (1.41–1.45) | 1.13 (1.11–1.16) | 1.14 (1.13, 1.16) | 1.42 (1.40, 1.43) |

Hazard ratios (HRs) with 95% confidence intervals (CIs) are presented for the associations of the electronic frailty index (eFI), Hospital Frailty Risk Score (HFRS), and Charlson Comorbidity Index (CCI) with all-cause mortality, severe infections, and fractures. For healthcare utilization, odds ratios (OR) indicate the likelihood of any healthcare utilization, while the incidence rate ratios (IRR) reflect the frequency of contacts among individuals with healthcare utilization. All models were adjusted for age and sex.

The odds of any healthcare utilization increased with frailty level (mild: OR 2·62, 95% CI 2·55–2·69; moderate: OR 3·56, 95% CI 3·41–3·72; severe: OR 3·15, 95% CI 2·96–3·35, continuous: OR 1·12, 95% CI 1·11–1·12) (Table 2). Among individuals with healthcare utilization, frailty was associated with greater contact intensity, with progressively higher rates for mild (IRR 1·91, 95% CI 1·87–1·96), moderate (IRR 2·85, 95% CI 2·76–2·94), and severe frailty (IRR 3·98, 95% CI 3·80–4·16) as well as when using the continuous eFI (IRR 1·08, 95% CI 1·08–1·08). The eFI was also significantly associated with increased odds of experiencing any event and higher conditional event rates of severe infections and fractures (Supplementary Table 7, appendix p 10).

The eFI-associated risks were similar in males and females (Supplementary Table 8, appendix p 11). In the age-group stratified analyses for the time-to-event outcomes and healthcare utilization, younger individuals exhibited relatively higher risks with increasing frailty (Supplementary Table 9, appendix p 12). When using the eFI as a continuous variable and restricting the analyses to individuals classified as non-frail (eFI ≤8), we observed significant associations with all outcomes (Table 3). Compared with the full sample, the estimates were slightly lower for all-cause mortality (HR 1·06 vs 1·09), but slightly higher for severe infections (HR 1·12 vs 1·10), fractures (HR 1·08 vs 1·05). and healthcare utilization for both the likelihood of utilization (OR 1·29 vs 1·12) and the frequency of healthcare contacts (IRR 1·11 vs 1·08) (Table 3).

**Table 3.** Associations between baseline eFI, and time-to-event and count outcomes in the non-frail category.

| Outcome | eFI baseline | Hazard/Odds/Incidence ratio (95% CI) | N | Events |
| --- | --- | --- | --- | --- |
| All-cause mortality | All | 1·06 (1·04 – 1·07) | 98677 | 3349 |
|  | Male | 1·06 (1·04 – 1·07) | 46108 | 1877 |
|  | Female | 1·06 (1·04 – 1·08) | 52569 | 1472 |
| Severe infections | All | 1·12 (1·11 – 1·14) | 106662 | 2499 |
|  | Male | 1·13 (1·11 – 1·16) | 50284 | 1222 |
|  | Female | 1·11 (1·09 – 1·13) | 56378 | 1277 |
| Fractures | All | 1·08 (1·07 – 1·09) | 106645 | 6075 |
|  | Male | 1·09 (1·07 – 1·10) | 50276 | 2506 |
|  | Female | 1·08 (1·07 – 1·10) | 56369 | 3569 |
| Healthcare utilization, odds | All | 1·29 (1·28 – 1·29) | 133810 | 62802 |
|  | Male | 1·26 (1·26 – 1·27) | 64573 | 27688 |
|  | Female | 1·31 (1·30 – 1·31) | 69237 | 35114 |
| Healthcare utilization, incidence rate | All | 1·11 (1·10 – 1·11) | 133810 | - |
|  | Male | 1·10 (1·09 – 1·11) | 64573 | - |
|  | Female | 1·11 (1·11 – 1·12) | 69237 | - |

Estimates are presented as hazard ratios (HRs) for mortality, severe infections, and fractures; odds ratios (ORs) and incidence rate ratios (IRRs) for healthcare use, with 95% confidence intervals (CIs) for the electronic frailty index (eFI). HRs were derived from Cox proportional hazards models, and ORs and IRRs from hurdle negative binomial models. The number of individuals (N) and number of events for each outcome are shown. All models were adjusted for baseline age and sex. n, number of individuals.

### Model performance

Addition of the eFI to the Cox base model (age and sex) either as a categorical or continuous variable resulted in the greatest improvement in model discrimination across all time-to-event outcomes compared with CCI and HFRS (Figure 3). For the count models (severe infections, fractures, and healthcare utilization), inclusion of eFI to the base model resulted in the greatest improvement in model fit compared with models including CCI and HFRS (Supplementary Tables 10–11, appendix pp 13–14).

Results on generalizability and model fit evaluation are presented in Supplementary Results (appendix p 1) and Supplementary Tables 12–14 (appendix pp 15–17).

## Discussion

In this large study based on EHR data from Central Finland, spanning primary, secondary and tertiary care, we developed a next-generation eFI based on routinely collected structured (diagnoses, laboratory tests) and unstructured (free-text) data. For the identification of the free-text eFI items, we leveraged a novel deep-learning AI-guided approach,^19^ that allowed us to capture frailty beyond what can be measured using conventional comorbidity-based or structured administrative data measures. Our eFI demonstrated robust predictive validity across all examined outcomes and across the adult age spectrum, including younger individuals, a population in which frailty has been understudied but where early vulnerability is nevertheless consequential. Importantly, the eFI achieved good resolution even at very low frailty levels, that is, among individuals classified as non-frail. The eFI also outperformed the HFRS and CCI across the outcomes, suggesting that it provides additional value for risk stratification over scales relying on ICD codes only.

The prevalence of moderate and severe frailty increased markedly with age; among individuals aged 35–49 years, only 4·1% were classified as moderately frail and 2·0% as severely frail, whereas in those aged ≥80 years, the corresponding proportions increased to 34·7% and 51·3%, respectively. Moderate frailty was present in 19·7% of individuals aged 50–64 years and 41·6% of those aged 65–79 years, while severe frailty increased from 11·6% to 35·1% across the same age categories. The apparently high prevalence of severe frailty should be interpreted with caution, as the eFI threshold used here to define it (>21 on the 0–100 scale) corresponds approximately with the conventional frailty cut-off commonly used in clinical assessment. The trajectory of the eFI accelerated after age 65, consistently with observations from cohort studies using the conventional FI.^20^

The frailty-associated risks were similar in males and females across all outcomes, whereas the relative risks associated with higher frailty were greater in younger age groups. These findings are consistent with cohort studies using the conventional FI, which similarly report greater relative risks of mortality in younger age groups.^21–23^ In addition, a previous study evaluating the UK eFI among younger adults aged 18–64 years demonstrated that the index predicts both 1- and 3-year mortality as well as emergency hospitalization.^24^ Most importantly, all our associations remained statistically significant when the analyses were restricted to individuals classified as non-frail, indicating that our eFI has good resolution to capture clinically relevant vulnerability even at very low levels of frailty. To our knowledge, such analyses specifically within a non-frail population have not previously been done and our findings provide novel insight into the ability of the eFI to identify vulnerability even below the lowest frailty threshold, hence supporting its use as a continuous measure among the non-frail.

Overall, our findings reinforce the dose-response relationship between frailty and the risk of death. This pattern is consistent with previous studies using eFI across healthcare settings, from the first primary care–based eFI,^10^ to more recent studies among hospitalized patients, all showing progressively higher mortality risk with increasing frailty levels.^13,25,26^ The comparatively stronger associations observed in our study may reflect the more comprehensive identification of deficits achieved by combining structured EHR data with items extracted from unstructured free-text records. In addition, the included laboratory tests may signal subclinical physiological changes that are not yet reflected in diagnostic codes. Therefore, our eFI approach may not only facilitate improved risk identification but also provide greater flexibility for developing transferable eFI solutions across healthcare systems and countries.

Our findings revealed that a higher eFI was associated with increased risk of severe infections up to two years from the eFI assessment, with large effect sizes (HR of 9·22 for severe frailty). In a study on the UK Biobank participants, higher FI levels were associated with progressively increased risk of severe infections, with HR reaching 3·37 in the highest FI category.^27^ A systematic review also reported that frailty is associated with infection risk, with the FI more often showing positive associations than phenotypic definitions of frailty.^28^ Our findings give support the use of the eFI to identify individuals at increased risk of severe, hospital-treatment– requiring infections, although its discrimination was lower than for all-cause mortality.

Higher eFI scores were also predictive of increased risk of fractures. This result is consistent with previous studies reporting progressively higher fracture risk across eFI categories,^29^ and with a recent meta-analysis showing an overall increased risk among frail subjects.^30^ Although and the discrimination of the eFI (0·62) for fracture risk was lower than that for mortality and fractures, it is nevertheless comparable to established fracture risk tools, such as FRAX, which has a C-index of ∼0·61.^31^ The eFI was also strongly and positively associated with healthcare utilization, showing a similar dose–response relationship across frailty categories similar to that observed for the other outcomes. This finding is consistent with previous studies showing increasing resource usage with greater frailty severity.^32,33^ Based on these results, the eFI could be used to support more proactive and better-aligned care planning in routine practice, helping clinicians identify individuals with increasing care needs and thus in need for earlier intervention, or preventive care approaches that may prevent avoidable hospitalizations.

The larger effect size for all-cause mortality when using the continuous eFI in the full sample vs the non-frail subgroup suggests that frailty contributes more strongly to mortality risk once deficits accumulate beyond the non-frail stage. In contrast, the stronger associations observed in the non-frail group for severe infections, fractures, and healthcare utilization suggest that the eFI also captures early physiological vulnerability that manifests through acute stressor-related outcomes rather than mortality. That is, subtle health deficits identified by the eFI may reflect already diminished resilience, and within a population considered relatively healthy, even small differences in vulnerability become clinically meaningful. Given that frailty is difficult to reverse but can be prevented or postponed, early detection of such subtle vulnerability may offer opportunities for targeted prevention and cost savings in healthcare.

This study has several strengths, including the use of longitudinal, large-scale, real-world EHR data spanning primary, secondary, and tertiary care within a tax-funded, universally accessible public healthcare system that minimizes socioeconomic barriers to care and ensures comprehensive population coverage. Our approach of constructing a multidimensional eFI incorporating diagnoses, routine laboratory measures, and free-text–derived information may offer an applicable method of measuring frailty across different healthcare systems. However, the study also has limitations, including possibly incomplete data capture inherent to all EHR-based research; the absence of direct lifestyle and socioeconomic measures due to collection limitations and privacy laws; and the observational design, which precludes causal inference.

## Conclusion

Our findings demonstrate that the EHR-based multidimensional eFI offers meaningful risk stratification across adulthood, capturing risks associated with both established frailty and early vulnerability. Its predictive performance across multiple outcomes, and added value over established risk scores, supports its potential as a scalable tool for proactive care planning.

## Data Availability

The authors cannot share the data due to legislative and ethical restrictions; however, all data generated in this study are available through an application to the Central Finland Wellbeing Services County.

## References

1. Clegg A, Young J, Iliffe S, et al. Frailty in elderly people. Lancet Lond Engl 2013; 381: 752–762.

2. Rockwood K, Song X, MacKnight C, et al. A global clinical measure of fitness and frailty in elderly people. CMAJ Can Med Assoc J J Assoc Medicale Can 2005; 173: 489–495.

3. Hoogendijk EO, Afilalo J, Ensrud KE, et al. Frailty: implications for clinical practice and public health. The Lancet 2019; 394: 1365–1375.

4. Kojima G, Iliffe S, Walters K. Frailty index as a predictor of mortality: a systematic review and meta-analysis. Age Ageing 2018; 47: 193–200.

5. Fried LP, Tangen CM, Walston J, et al. Frailty in Older Adults: Evidence for a Phenotype. J Gerontol A Biol Sci Med Sci 2001; 56: M146–M157.

6. Best K, Shuweihdi F, Alvarez JCB, et al. Development and external validation of the electronic frailty index 2 using routine primary care electronic health record data. Age Ageing 2025; 54: afaf077.

7. Zheng J, Yu P, Yang M. Development, Validation, and Application of the Electronic Frailty Index: A Scoping Review. J Am Med Dir Assoc 2025; 26: 105577.

8. Khanna AK, Motamedi V, Bouldin B, et al. Automated Electronic Frailty Index–Identified Frailty Status and Associated Postsurgical Adverse Events. JAMA Netw Open 2023; 6: e2341915.

9. Fujita K, Lo SY, Hubbard RE, et al. Comparison of a multidomain frailty index from routine health data with the hospital frailty risk score in older patients in an Australian hospital. Australas J Ageing 2023; 42: 480–490.

10. Mak JKL, Hägg S, Eriksdotter M, et al. Development of an Electronic Frailty Index for Hospitalized Older Adults in Sweden. J Gerontol Ser A 2022; 77: 2311–2319.

11. Bai G, Wang Y, Mak JKL, et al. Is Frailty Different in Younger Adults Compared to Old? Prevalence, Characteristics, and Risk Factors of Early-Life and Late-Life Frailty in Samples from Sweden and UK. Gerontology 2023; 69: 1385–1393.

12. Morales DR, Guthrie B, Downes TJ, et al. Applicability of the electronic frailty index in younger and older adults in England: a population-based cohort study. Lancet Healthy Longev 2025; 6: 100752.

13. Clegg A, Bates C, Young J, et al. Development and validation of an electronic frailty index using routine primary care electronic health record data. Age Ageing 2016; 45: 353–360.

14. Bai C, Mardini MT. Advances of artificial intelligence in predicting frailty using real-world data: A scoping review. Ageing Res Rev 2024; 101: 102529.

15. Plasek JM, Leeson M, Lundstedt J, et al. Validation of a Large Language Model Enhanced Frailty Index. J Med Syst 2026; 50: 53.

16. The Wellbeing Services County of Central Finland. Keski-Suomen hyvinvointialue, https://www.hyvaks.fi/sites/default/files/2025-07/The%20wellbeing%20services%20county%20of%20Central%20Finland%20_04072025.pdf (4 July 2025, accessed 15 June 2026).

17. Searle SD, Mitnitski A, Gahbauer EA, et al. A standard procedure for creating a frailty index. BMC Geriatr 2008; 8: 24.

18. Cybularz M, Wydra S, Berndt K, et al. Frailty is associated with chronic inflammation and pro-inflammatory monocyte subpopulations. Eur Heart J 2020; 41: ehaa946.1881.

19. Lin J, Kuukka A, Korpi T, et al. Identifying health conditions in older adults in textual health records using deep learning-based natural language processing. Comput Struct Biotechnol J 2025; 28: 341–347.

20. Raymond E, Reynolds CA, Dahl Aslan AK, et al. Drivers of Frailty from Adulthood into Old Age: Results from a 27-Year Longitudinal Population-Based Study in Sweden. J Gerontol A Biol Sci Med Sci 2020; 75: 1943–1950.

21. Fan J, Yu C, Guo Y, et al. Frailty index and all-cause and cause-specific mortality in Chinese adults: a prospective cohort study. Lancet Public Health 2020; 5: e650–e660.

22. Mak JKL, Kuja-Halkola R, Wang Y, et al. Frailty and comorbidity in predicting community COVID-19 mortality in the UK Biobank: the effect of sampling. Epub ahead of print 27 October 2020. DOI: 10.1101/2020.10.22.20217489.

23. Li X, Ploner A, Karlsson IK, et al. The frailty index is a predictor of cause-specific mortality independent of familial effects from midlife onwards: a large cohort study. BMC Med 2019; 17: 94.

24. Morales DR, Guthrie B, Downes TJ, et al. Applicability of the electronic frailty index in younger and older adults in England: a population-based cohort study. Lancet Healthy Longev 2025; 6: 100752.

25. Liang Y-D, Xie Y-B, Du M-H, et al. Development and Validation of an Electronic Frailty Index Using Routine Electronic Health Records: An Observational Study From a General Hospital in China. Front Med 2021; 8: 731445.

26. Lowry MTH, Kimenai DM, Doudesis D, et al. The electronic frailty index and outcomes in patients with myocardial infarction. Age Ageing 2024; 53: afae150.

27. Bai R, Pan L, Huang Y, et al. Association of accelerated biological aging and frailty with the risk of severe infection: a prospective study in the UK Biobank. J Frailty Aging 2026; 15: 100118.

28. Cosentino CB, Mitchell BG, Brewster DJ, et al. The utility of frailty indices in predicting the risk of health care associated infections: A systematic review. Am J Infect Control 2021; 49: 1078–1084.

29. Middleton R, Poveda JL, Orfila Pernas F, et al. Mortality, Falls, and Fracture Risk Are Positively Associated With Frailty: A SIDIAP Cohort Study of 890 000 Patients. J Gerontol Ser A 2022; 77: 148–154.

30. Kojima G. Prevalence of Frailty in Nursing Homes: A Systematic Review and Meta-Analysis. J Am Med Dir Assoc 2015; 16: 940–945.

31. Li G, Thabane L, Papaioannou A, et al. Comparison between frailty index of deficit accumulation and fracture risk assessment tool (FRAX) in prediction of risk of fractures. Bone 2015; 77: 107–114.

32. Han L, Clegg A, Doran T, et al. The impact of frailty on healthcare resource use: a longitudinal analysis using the Clinical Practice Research Datalink in England. Age Ageing 2019; 48: 665–671.

33. Ikonen JN, Eriksson JG, Salonen MK, et al. The utilization of specialized healthcare services among frail older adults in the Helsinki Birth Cohort Study. Ann Med 2021; 53: 1875–1884.

