## Supplementary data for "A next-generation electronic frailty index leveraging deep learning on unstructured health records extends risk prediction across the full frailty spectrum"

### Supplementary appendix


| | |
|---|---|
| Page 1 | Supplementary Methods |
| Page 1 | Supplementary Results |
| Page 2 | Supplementary References |
| Page 3 | Supplementary Table 1. Items included in the eFI. |
| Page 5 | Supplementary Table 2. ICD-10 codes used to define outcomes. |
| Page 6 | Supplementary Table 3. Baseline characteristics of the population stratified by eFI categories. |
| Page 7 | Supplementary Table 4. Performance metrics of the deficits identified from free-text items using NLP NER. |
| Page 8 | Supplementary Table 5. Correlation coefficients among the eFI items at baseline. |
| Page 9 | Supplementary Table 6. Generalized additive mixed model for the eFI trajectories. |
| Page 10 | Supplementary Table 7. Associations between eFI and severe infections and fractures in the count models. |
| Page 11 | Supplementary Table 8. Associations between eFI and time-to-event and count outcomes across male and female. |
| Page 12 | Supplementary Table 9. Associations between eFI and time-to-event and count outcomes across age groups. |
| Page 13 | Supplementary Table 10. Model fit statistics for severe infections and fractures in count models. |
| Page 14 | Supplementary Table 11. Model fit statistics for examining total healthcare utilization. |
| Page 15 | Supplementary Table 12. Hold-out validation based on model concordance for models on all-cause mortality, severe infections, and fractures. |
| Page 16 | Supplementary Table 13. Cross-validated performance for severe infections and fractures based on count models. |
| Page 17 | Supplementary Table 14. Cross-validated model performance for the total healthcare u models. |
| Page 18 | Supplementary Figure 1. Flowchart of baseline eFI construction. |
| Page 19 | Supplementary Figure 2. Study design and time windows for baseline eFI assessment and outcome follow-up. |
| Page 20 | Supplementary Figure 3. Outcome-specific analytical samples derived from the baseline eFI cohort. |
| Page 21 | Supplementary Figure 4. Kaplan–Meier curves for all-cause mortality, severe infections, and fractures by eFI categories. |

### Supplementary Methods

#### *Baseline eFI Calculation*

Baseline eFI was calculated using data from the first three years after each individual's initial EHR entry, with the baseline value defined at the end of this three-year period. For example, subjects entering the EHR in 2012 had their baseline eFI calculated using data from 2012, 2013, and 2014. All unique deficits observed during this three-year baseline period were summed. The baseline eFI was calculated as the ratio of the number of observed deficits to a person- and time -specific denominator representing the number of eligible deficits for that individual in that particular year and multiplied by a 100 to facilitate interpretation. Because laboratory test availability varied across subjects, denominators ranged from 44 to 53. The eFI was thus defined as:

$$\text{eFI} = \frac{\text{Number of observed deficits}}{\text{Total number of available deficits}} * 100$$

In addition to treating the baseline eFI as a continuous variable, we categorized it using data-driven thresholds based on observed mortality risk. Specifically, the number of categories and optimal cut points were identified using maximally selected log-rank statistics based on Kaplan–Meier survival curves for all-cause mortality, subject to predefined minimum group-size constraints. This procedure was applied iteratively to identify additional data-driven cut-offs within higher eFI ranges.

#### Calculation of HFRS and CCI

HFRS and CCI were calculated using ICD-10 codes recorded during the same three-year baseline period as the eFI, and similar to the eFI, considered as both categorical and continuous measures. HFRS was categorized into the established risk groups (low <5, intermediate 5–15, high >15), as described by Gilbert and colleagues.[1] CCI was calculated using the standard algorithm[2] and its updated ICD-10–based mapping,[3] and categorized as follows: no comorbidity (0), mild comorbidity (1), moderate comorbidity (2), and severe comorbidity (≥3).

#### Statistical Analysis of Count Models

Counts of severe infections, fracture episodes, and healthcare utilization were analysed using hurdle negative binomial models. A hurdle model estimates the likelihood of any healthcare utilization and the frequency of contacts among individuals with healthcare utilization; the binomial component produced odds ratios (ORs) for any use, while the truncated count component produced incidence rate ratios (IRRs) reflecting contact intensity. Model fit for count models was assessed using the Akaike Information Criterion (AIC) and Bayesian Information Criterion (BIC). Generalizability of the count models was evaluated using a 70:30 train–test split with 10-fold cross-validation, with root mean squared error (RMSE) and mean absolute error (MAE) used to quantify prediction error.

### Supplementary Results

For the time-to-event outcomes, hold-out validation demonstrated highly consistent results between the training and test sets across the models (Supplementary Table 12, appendix p 15), indicating that the model generalized well to unseen data and was not dependent on the sample used for model development.

In the count models (severe infections, fractures, and healthcare use), RMSE and MAE were consistent between training and hold-out test sets, indicating stable out-of-sample performance. Models including eFI achieved the lowest RMSE and MAE, reflecting modest improvements in predictive accuracy compared with the base model, whereas CCI and HFRS showed more limited gains (Supplementary Table 13–14, appendix pp 16–17).

**Supplementary References**

1. Gilbert T, Neuburger J, Kraindler J, et al. Development and validation of a Hospital Frailty Risk Score focusing on older people in acute care settings using electronic hospital records: an observational study. *The Lancet* 2018; 391: 1775–1782.

2. Charlson ME, Pompei P, Ales KL, et al. A new method of classifying prognostic comorbidity in longitudinal studies: Development and validation. *J Chronic Dis* 1987; 40: 373–383.

3. Ludvigsson JF, Appelros P, Askling J, et al. Adaptation of the Charlson Comorbidity Index for Register-Based Research in Sweden. *Clin Epidemiol* 2021; 13: 21–41.

**Supplementary Table 1. Items included in the eFI.**

Deficits were classified as either permanent or resettable. Laboratory and free-text–derived deficits were treated as resettable and carried forward for up to 2 years after the last abnormal (outside reference range) measurement or recorded condition, after which they were reset if not re-observed. Diagnosis-based deficits were considered permanent once recorded, except for five predefined resettable items.

| No. | Deficit | Coding |
|---|---|---|
| 1 | Asthma | 1 = Any of the ICD-10 codes: J45, J46 |
| 2 | Atrial fibrillation | 1 = Any of the ICD-10 codes: I48 |
| 3 | Cancer | 1 = Any of the ICD-10 codes: C00-C97 |
| 4 | Cardiac valve disease | 1 = Any of the ICD-10 codes: I05-I08, I09·1, I09·8, I34-I39, Z95·2-Z95·4 |
| 5 | Cerebrovascular disease | 1 = Any of the ICD-10 codes: I60-I66 |
| 6 | Chronic liver disease | 1 = Any of the ICD-10 codes: K70-K76, Z94·4 |
| 7 | Connective tissue diseases | 1 = Any of the ICD-10 codes: L10, L12, L40, L41, L93-L95, M30-M36 |
| 8 | COPD, chronic bronchitis, and emphysema | 1 = Any of the ICD-10 codes: J41-J44, J47 |
| 9 | Dementia | 1 = Any of the ICD-10 codes: F00-F03, F05·1, G30, G31 |
| 10 | Diabetes | 1 = Any of the ICD-10 codes: E10-E14 |
| 11 | Diseases of the gastrointestinal tract | 1 = Any of the ICD-10 codes: K80-K87, K90·0 |
| 12 | Diseases of the urinary tract | 1 = Any of the ICD-10 codes: N20-N21, N23, N31-N32, N35-N36, N39·1-N39·9 |
| 13 | Dizziness or vertigo* | 1 = Any of the ICD-10 codes: H81, R42 |
| 14 | Dyspnoea* | 1 = Any of the ICD-10 codes: R06·0 |
| 15 | Epilepsy | 1 = Any of the ICD-10 codes: G40, G41 |
| 16 | Gallbladder, bile duct, and pancreatic diseases | 1 = Any of the ICD-10 codes: K20-K31, K35-K38, K40-K46, K50-K52, K55-K63 |
| 17 | Heart failure | 1 = Any of the ICD-10 codes: I11·0, I13·0, I13·2, I27, I28·0, I42, I43, I50, I51·5, I51·7, Z94·1, Z94·3 |
| 18 | Hypertension | 1 = Any of the ICD-10 codes: I10, I15 |
| 19 | Inflammatory arthropathies | 1 = Any of the ICD-10 codes: M05-M07, M10-M13 |
| 20 | Ischemic heart disease | 1 = Any of the ICD-10 codes: I20, I21, I22, I24, I25, Z95·1, Z95·5 |
| 21 | Mood disorders | 1 = Any of the ICD-10 codes: F30-F39, F41 |
| 22 | Obesity | 1 = Any of the ICD-10 codes: E66 |
| 23 | Orthostatic hypotension* | 1 = Any of the ICD-10 codes: I95 |
| 24 | Osteoarthritis | 1 = Any of the ICD-10 codes: M15-M19 |
| 25 | Osteoporosis | 1 = Any of the ICD-10 codes: M80-M82 |
| 26 | Pain* | 1 = Any of the ICD-10 codes: G43, G44, G50·0, G50·1, G53·0, G54·6, F45·4, H57·1, H92·0, K07·63, K13·6, K14·6, M25·5, M54·2-M54·9, M77·4, M79·1, M79·2, M79·6, M79·7, M94·0, R07, R10, R51, R52 |
| 27 | Parkinson's disease and parkinsonism | 1 = Any of the ICD-10 codes: G20-G23 |
| 28 | Peripheral artery disease | 1 = Any of the ICD-10 codes: I70·2 |
| 29 | Peripheral venous insufficiency | 1 = Any of the ICD-10 codes: I83, I87·2 |
| 30 | Pressure ulcer* | 1 = Any of the ICD-10 codes: L89 |
| 31 | Renal disease | 1 = Any of the ICD-10 codes: I12·0, I13·1, I13·2, I13·9, N00-N08, N10-N13, N17-N19, N25-N27, N28·0, N28·1, Z94·0 |
| 32 | Schizophrenia and delusional disorders | 1 = Any of the ICD-10 codes: F20, F22, F24, F25, F28 |
| 33 | Sleep disorders | 1 = Any of the ICD-10 codes: F51, G47 |
| 34 | Thyroid disease | 1 = Any of the ICD-10 codes: E00-E07, E89·0 |

| | | |
|---|---|---|
| 35 | Mobility limitations*§ | 0 = No problem, 1 = Problem present |
| 36 | Visual impairment*§ | 0 = No problem, 1 = Problem present |
| 37 | Hearing impairment*§ | 0 = No problem, 1 = Problem present |
| 38 | Needing help with bathing*§ | 0 = No problem, 1 = Problem present |
| 39 | Needing help with dressing*§ | 0 = No problem, 1 = Problem present |
| 40 | Needing help with eating and/or preparing meals*§ | 0 = No problem, 1 = Problem present |
| 41 | Incontinence*§ | 0 = No problem, 1 = Problem present |
| 42 | Falls*§ | 0 = No problem, 1 = Problem present |
| 43 | Age-related neurocognitive problems*§ | 0 = No problem, 1 = Problem present |
| 44 | Loneliness*§ | 0 = No problem, 1 = Problem present |
| 45 | HbA1c* | 1 = HbA1c > 42 mmol/mol |
| 46 | Haemoglobin* | 1 = Haemoglobin < 117 g/L (women) or < 134 g/L (men) |
| 47 | Potassium* | 1 = Potassium < 3·3 mmol/L or > 4·9 mmol/L |
| 48 | Sodium* | 1 = Sodium < 137 mmol/L or > 145 mmol/L |
| 49 | Triglycerides* | 1 = Triglycerides > 1·7 mmol/L |
| 50 | LDL cholesterol* | 1 = LDL cholesterol > 3·0 mmol/L |
| 51 | C-reactive protein* | 1 = C-reactive protein > 4 mg/L |
| 52 | Creatinine* | 1 = Creatinine < 50 µmol/L or > 90 µmol/L (women) or < 60 µmol/L or > 100 µmol/L (men) |
| 53 | Glucose* | 1 = Fasting plasma glucose < 4·0 mmol/L or > 6·0 mmol/L (non-fasting values excluded) |

*Reset after two years.
§Identified from the free texts using AI-driven NLP
COPD, chronic obstructive pulmonary disease; eFI, electronic Frailty Index; HbA1c, glycated haemoglobin; ICD-10, International Classification of Diseases, 10th Revision; LDL, low-density lipoprotein cholesterol; NLP, Natural Language Processing.

**Supplementary Table 2. ICD-10 codes used to define severe hospital treatment requiring infections and fractures.**

| Outcome | ICD-10 codes |
|---|---|
| Severe infections | A40–A41; G00–G04; G06; I33; J12–J18; M00–M01, M46, M72·6, M86; N10–N12; K65; A04·7, A48·0; J86; T81·4, T84·5, T84·7, T85·7 |
| Fractures | S02, S12, S22, S32, S42, S52, S62, S72, S82, S92; T02, T10, T12 |

**Supplementary Table 3. Baseline characteristics of the population stratified by eFI categories.**

| Characteristic | Overall<br>N = 193629 | Non-frail<br>N = 149090 | Mild Frailty<br>N = 27808 | Moderate Frailty<br>N = 11471 | Severe Frailty<br>N = 5260 | p-value |
|---|---|---|---|---|---|---|
| **eFI** | | | | | | <0·0001 |
| Mean ± SD | 4·9 ± 6·2 | 2·1 ± 2·4 | 10·8 ± 1·8 | 17·4 ± 1·9 | 25·8 ± 4·1 | |
| Median (Q1, Q3) | 2·3 (0·0, 7·7) | 2·0 (0·0, 4·0) | 10·2 (9·4, 12·2) | 17·0 (15·7, 18·9) | 24·5 (22·6, 28·3) | |
| **Age** | | | | | | <0·0001 |
| Mean ± SD | 62·0 ± 13·6 | 59·4 ± 12·7 | 68·3 ± 13·0 | 73·0 ± 12·1 | 77·4 ± 11·1 | |
| Median (Q1, Q3) | 62·0 (51·0, 72·0) | 59·0 (49·0, 68·0) | 68·0 (59·0, 78·0) | 74·0 (65·0, 83·0) | 80·0 (70·0, 86·0) | |
| **HFRS** | | | | | | <0·0001 |
| Mean ± SD | 0·8 ± 1·9 | 0·4 ± 1·2 | 1·4 ± 2·3 | 2·6 ± 3·2 | 4·7 ± 4·5 | |
| Median (Q1, Q3) | 0·0 (0·0, 0·9) | 0·0 (0·0, 0·0) | 0·5 (0·0, 2·1) | 1·6 (0·0, 3·6) | 3·5 (1·5, 6·5) | |
| **CCI** | | | | | | <0·0001 |
| Mean ± SD | 0·3 ± 0·9 | 0·1 ± 0·5 | 0·7 ± 1·2 | 1·2 ± 1·4 | 2·0 ± 1·7 | |
| Median (Q1, Q3) | 0·0 (0·0, 0·0) | 0·0 (0·0, 0·0) | 0·0 (0·0, 1·0) | 1·0 (0·0, 2·0) | 2·0 (1·0, 3·0) | |
| **Gender**, N (%) | | | | | | <0·0001 |
| Female | 99837 (51·6) | 77064 (51·7) | 13973 (50·2) | 5910 (51·5) | 2890 (54·9) | |
| Male | 93792 (48·4) | 72026 (48·3) | 13835 (49·8) | 5561 (48·5) | 2370 (45·1) | |
| **Age group**, N (%) | | | | | | <0·0001 |
| 35–49 | 40858 (21·1) | 37750 (25·3) | 2537 (9·1) | 467 (4·1) | 104 (2·0) | |
| 50–64 | 70807 (36·6) | 60079 (40·3) | 7860 (28·3) | 2258 (19·7) | 610 (11·6) | |
| 65–79 | 58316 (30·1) | 40472 (27·1) | 11225 (40·4) | 4771 (41·6) | 1848 (35·1) | |
| ≥80 | 23648 (12·2) | 10789 (7·2) | 6186 (22·2) | 3975 (34·7) | 2698 (51·3) | |
| **HFRS Categories**, N (%) | | | | | | <0·0001 |
| Low risk (<5) | 186314 (96·2) | 147354 (98·8) | 25939 (93·3) | 9614 (83·8) | 3407 (64·8) | |
| Intermediate risk (5–15) | 6923 (3·6) | 1716 (1·2) | 1798 (6·5) | 1754 (15·3) | 1655 (31·5) | |
| High risk (>15) | 392 (0·2) | 20 (0·0) | 71 (0·3) | 103 (0·9) | 198 (3·8) | |
| **CCI categories**, N (%) | | | | | | <0·0001 |
| No comorbidity (=0) | 157670 (81·4) | 135731 (91·0) | 16547 (59·5) | 4460 (38·9) | 932 (17·7) | |
| Mild comorbidity (=1) | 18246 (9·4) | 7792 (5·2) | 5835 (21·0) | 3144 (27·4) | 1475 (28·0) | |
| Moderate comorbidity (=2) | 12928 (6·7) | 5090 (3·4) | 4181 (15·0) | 2414 (21·0) | 1243 (23·6) | |
| Severe comorbidity (≥3) | 4785 (2·5) | 477 (0·3) | 1245 (4·5) | 1453 (12·7) | 1610 (30·6) | |
| **Death,** N (%) | | | | | | <0·0001 |
| No | 155023 (80·1) | 132650 (89·0) | 16909 (60·8) | 4500 (39·2) | 964 (18·3) | |
| Yes | 38606 (19·9) | 16440 (11·0) | 10899 (39·2) | 6971 (60·8) | 4296 (81·7) | |
| **Severe infections**, N (%) | | | | | | <0·0001 |
| No | 187136 (96·6) | 146591 (98·3) | 26215 (94·3) | 10132 (88·3) | 4198 (79·8) | |
| Yes | 6493 (3·4) | 2499 (1·7) | 1593 (5·7) | 1339 (11·7) | 1062 (20·2) | |
| **Fractures**, N (%) | | | | | | <0·0001 |
| No | 183754 (94·9) | 143015 (95·9) | 25755 (92·6) | 10325 (90·0) | 4659 (88·6) | |
| Yes | 9875 (5·1) | 6075 (4·1) | 2053 (7·4) | 1146 (10·0) | 601 (11·4) | |
| **Healthcare Use**, N (%) | | | | | | <0·0001 |
| No | 100221 (51·8) | 86288 (57·9) | 9169 (33·0) | 3156 (27·5) | 1608 (30·6) | |
| Yes | 93408 (48·2) | 62802 (42·1) | 18639 (67·0) | 8315 (72·5) | 3652 (69·4) | |

Baseline characteristics of the study population stratified by electronic frailty index (eFI) categories. Characteristics are presented overall and by eFI category (non-frail, mild frailty, moderate frailty, and severe frailty). Continuous variables are summarized as mean ± SD and median (Q1, Q3), and categorical variables as n (%), where n denotes the number of individuals and % denotes the proportion within each group. For binary variables such as severe infections, fractures, and healthcare use, Yes and No indicate presence and absence of the characteristic in the defined observation period. P values compare differences across eFI categories using the Kruskal–Wallis rank-sum test for continuous variables and Pearson's $\chi^2$ test for categorical variables.

CCI, Charlson Comorbidity Index; eFI, electronic frailty index; HFRS, Hospital Frailty Risk Score.

**Supplementary Table 4. Performance metrics of the deficits identified from free-text items using NLP NER.**

| Category | F1 | Precision | Recall | Evaluation type (strictness) | Learning rate | Weight decay |
|---|---|---|---|---|---|---|
| Falls | 0·87 | 0·88 | 0·86 | partial | 1·0E-05 | 0·20000 |
| Incontinence | 0·81 | 0·78 | 0·84 | partial | 1·8E-04 | 0·20003 |
| Loneliness | 0·88 | 0·86 | 0·91 | partial | 1·0E-04 | 0·19997 |
| Mobility limitations | 0·85 | 0·84 | 0·86 | partial | 2·0E-05 | 0·20000 |
| Hearing impairment | 0·79 | 0·75 | 0·82 | ent_type | 3·0E-05 | 0·19993 |
| Visual impairment | 0·82 | 0·70 | 0·98 | ent_type | 5·0E-06 | 0·01000 |
| Needs help with bathing | 0·92 | 0·89 | 0·96 | ent_type | 8·0E-05 | 0·19995 |
| Needs help with dressing | 0·86 | 0·77 | 0·98 | ent_type | 2·4E-05 | 0·19995 |
| Needs help with eating or preparing meals | 0·74 | 0·71 | 0·78 | ent_type | 9·0E-05 | 0·19996 |
| Age-related neurocognitive problems | 0·91 | 0·86 | 0·96 | ent_type | 3·5E-05 | 0·24000 |

Precision represents the accuracy of positive predictions; Recall indicates the ability to identify all actual positive instances; the F1 score provides the harmonic mean of both precision and recall, serving as a balanced measure of model reliability. Evaluation type defines what counts as a correct prediction relative to the ground truth; partial evaluation requires that predicted and ground truth spans overlap, regardless of entity type. The ent_type evaluation additionally requires a correct entity type but does not require exact boundary matches. The learning rate controls the size of each weight update during training. Weight decay is a regularization technique that reduces large weights. NLP, natural language processing, NER, named entity recognition

**Supplementary Table 5. Correlation coefficients among the eFI items at baseline.**

| Table | Variable 1 | Variable 2 | Correlation |
|---|---|---|---|
| Top 10 correlations across all items | Fasting glucose | Glycated haemoglobin | 0·561 |
| | Haemoglobin | C-reactive protein | 0·551 |
| | Sodium | C-reactive protein | 0·469 |
| | Sodium | Haemoglobin | 0·464 |
| | Glycated haemoglobin | Diabetes | 0·442 |
| | Sodium | Potassium | 0·434 |
| | Dressing | Bathing | 0·399 |
| | Mobility | Bathing | 0·398 |
| | Potassium | Haemoglobin | 0·386 |
| | Creatinine | Haemoglobin | 0·384 |
| Top 3 correlations across laboratory tests | Fasting glucose | Glycated haemoglobin | 0·561 |
| | Haemoglobin | C-reactive protein | 0·551 |
| | Sodium | C-reactive protein | 0·469 |
| Top 3 correlations across ICD-10 diagnoses | Heart failure | Atrial fibrillation | 0·245 |
| | COPD | Asthma | 0·245 |
| | Hypertension | Diabetes | 0·218 |
| Top 3 correlations across free-text identified items | Dressing | Bathing | 0·399 |
| | Mobility | Bathing | 0·398 |
| | Feeding | Bathing | 0·350 |

The table presents the strongest correlations overall and within domains, including laboratory variables, diagnostic codes based on the International Classification of Diseases, 10th Revision (ICD-10), and items extracted from free-text clinical notes using natural language processing with named entity recognition.
COPD, chronic obstructive pulmonary disease.

**Supplementary Table 6. Generalized additive mixed model for the eFI trajectories.**
Results from a generalized additive model (GAM) with a Tweedie distribution and log link describing eFI trajectories over time. Parametric terms are presented as exponentiated coefficients with 95% confidence intervals (CIs). Smooth terms are reported as estimated degrees of freedom (EDF) with corresponding p values. Age_C denotes age centred around the sample mean age, and ID denotes the participant identifier used to specify subject-level random effects. Model fit is summarized by adjusted R² and deviance explained.

| **Parametric terms** | | |
|---|---|---|
| **Variable** | **exp(β) (95% CI)** | **p value** |
| Intercept | 6·584 (6·580–6·588) | <0·0001 |
| Male sex | 1·008 (1·004–1·011) | <0·0001 |
| **Smooth terms** | | |
| **Term** | **EDF** | **p value** |
| s(Age_C) – Female | 8·765 | <0·0001 |
| s(Age_C) – Male | 8·231 | <0·0001 |
| s(ID) | 0·992 | <0·0001 |
| s(ID, Age_C) | 0·937 | <0·0001 |
| **Model fit** | | |
| **Metric** | **Value** | |
| Adjusted $R^2$ | 0·241 | |
| Deviance explained | 13·8% | |

**Supplementary Table 7. Associations between eFI and severe infections and fractures in the count models.**

| | Severe Infections (N=142229) | | Fractures (N = 142209) | |
|---|---|---|---|---|
| | OR (95% CI) | IRR (95% CI) | OR (95% CI) | IRR (95% CI) |
| **eFI** | | | | |
| Categorical | | | | |
| Non-frail (≤8, Ref.) | 1 | 1 | 1 | 1 |
| Mild Frailty (>8–15) | 2·51 (2·35, 2·69) | 1·24 (1·10, 1·39) | 1·56 (1·48, 1·65) | 1·19 (1·07, 1·32) |
| Moderate Frailty (>15–21) | 4·94 (4·58, 5·32) | 1·49 (1·32, 1·69) | 2·12 (1·97, 2·27) | 1·46 (1·28, 1·67) |
| Severe Frailty (>21) | 9·78 (8·96, 10·68) | 1·85 (1·61, 2·12) | 2·60 (2·36, 2·86) | 1·72 (1·44, 2·05) |
| Continuous per point increase | 1·11 (1·10, 1·11) | 1·03 (1·02, 1·03) | 1·05 (1·05, 1·05) | 1·02 (1·01, 1·03) |
| **HFRS** | | | | |
| Categorical | | | | |
| Low risk (<5, Ref.) | 1 | 1 | 1 | 1 |
| Intermediate risk (5–15) | 2·91 (2·69, 3·15) | 1·38 (1·21, 1·58) | 1·84 (1·70, 2·00) | 1·34 (1·15, 1·56) |
| High risk (>15) | 3·96 (3·05, 5·14) | 0·74 (0·49, 1·12) | 2·58 (1·94, 3·42) | 1·58 (0·95, 2·65) |
| Continuous per point increase | 1·15 (1·14, 1·16) | 1·02 (1·00, 1·03) | 1·09 (1·09, 1·10) | 1·04 (1·02, 1·06) |
| **CCI** | | | | |
| Categorical | | | | |
| No comorbidity (=0) | 1 | 1 | 1 | 1 |
| Mild comorbidity (=1) | 2·46 (2·30, 2·62) | 1·14 (1·01, 1·27) | 1·13 (1·00, 1·27) | 1·13 (1·00, 1·27) |
| Moderate comorbidity (=2) | 2·78 (2·58, 3·00) | 1·40 (1·22, 1·60) | 1·27 (1·08, 1·48) | 1·27 (1·08, 1·48) |
| Severe comorbidity (≥3) | 5·13 (4·65, 5·66) | 1·45 (1·24, 1·71) | 1·16 (0·93, 1·45) | 1·16 (0·93, 1·45) |
| Continuous per point increase | 1·48 (1·45, 1·50) | 1·12 (1·07, 1·16) | 1·11 (1·09, 1·14) | 1·06 (1·01, 1·12) |

Hurdle negative binomial model estimates for severe infection and fracture counts. Odds ratios (ORs) and incidence rate ratios (IRRs) with 95% confidence intervals (CIs) are presented for the electronic frailty index (eFI), Hospital Frailty Risk Score (HFRS), and Charlson Comorbidity Index (CCI), modelled separately and adjusted for age and sex. Results are shown for both the hurdle (zero) component (OR) and the count component (IRR) of the models. In this context, the OR represents the odds of having at least one event (i.e., crossing the hurdle) versus having no events, whereas the IRR represents the relative rate of events among individuals with ≥1 event.

**Supplementary Table 8. Associations between eFI and time-to-event and count outcomes by sex.**

| | | All-cause mortality HR (95% CI) | Severe infections HR (95% CI) | Fractures HR (95% CI) | Healthcare utilization OR (95% CI) | Healthcare utilization IRR (95% CI) |
|---|---|---|---|---|---|---|
| Female | Mild Frailty | 2·12 (1·96–2·29) | 2·43 (2·21–2·67) | 1·59 (1·48–1·70) | 2·80 (2·69, 2·92) | 1·97 (1·92, 2·03) |
| Male | Mild Frailty | 2·16 (2·01–2·32) | 2·53 (2·31–2·77) | 1·51 (1·39–1·64) | 2·45 (2·36, 2·55) | 1·84 (1·78, 1·91) |
| Female | Moderate Frailty | 3·63 (3·33–3·96) | 4·76 (4·30–5·27) | 2·05 (1·88–2·24) | 3·70 (3·47, 3·94) | 2·78 (2·67, 2·90) |
| Male | Moderate Frailty | 4·24 (3·91–4·59) | 4·76 (4·31–5·25) | 2·19 (1·97–2·43) | 3·44 (3·24, 3·66) | 2·94 (2·81, 3·08) |
| Female | Severe Frailty | 6·80 (6·18–7·49) | 9·72 (8·70–10·86) | 2·69 (2·41–3·01) | 3·31 (3·04, 3·60) | 3·85 (3·64, 4·08) |
| Male | Severe Frailty | 7·61 (6·89–8·40) | 8·85 (7·90–9·92) | 2·81 (2·43–3·25) | 3·02 (2·76, 3·31) | 4·16 (3·88, 4·47) |
| Female | eFI continuous | 1·09 (1·08–1·09) | 1·10 (1·10–1·11) | 1·05 (1·05–1·05) | 1·12 (1·12, 1·13) | 1·08 (1·08, 1·08) |
| Male | eFI continuous | 1·10 (1·09–1·10) | 1·10 (1·10–1·11) | 1·05 (1·05–1·06) | 1·11 (1·11, 1·11) | 1·08 (1·08, 1·08) |

Estimates are presented as hazard ratios (HRs), odds ratios (ORs), and incidence rate ratios (IRRs) with 95% confidence intervals (CIs) for the electronic frailty index (eFI). HRs were derived from Cox proportional hazards models, and ORs and IRRs from hurdle negative binomial models. All models were adjusted for baseline age. For categorical eFI predictors, estimates are relative to the non-frail within each gender.

**Supplementary Table 9. Associations between eFI and time-to-event and count outcomes across age groups.**

| Age group | Predictor | Mortality HR (95% CI) | Severe Infections HR (95% CI) | Fractures HR (95% CI) | Healthcare use OR (95% CI) | Healthcare use IRR (95% CI) |
|---|---|---|---|---|---|---|
| 35–49 | Mild Frailty | 4·34 (3·28–5·75) | 3·28 (2·67–4·04) | 1·82 (1·56–2·11) | 3·70 (3·36, 4·08) | 2·70 (2·52, 2·90) |
| | Moderate Frailty | 12·17 (8·45–17·53) | 7·84 (5·80–10·59) | 2·21 (1·63–2·99) | 7·44 (5·65, 9·80) | 5·11 (4·40, 5·93) |
| | Severe Frailty | 38·55 (23·77–62·54) | 18·64 (12·14–28·62) | 3·10 (1·76–5·47) | 9·29 (4·96, 17·39) | 10·56 (7·73, 14·43) |
| | eFI continuous | 1·18 (1·16–1·20) | 1·15 (1·13–1·16) | 1·07 (1·06–1·08) | 1·20 (1·19, 1·21) | 1·14 (1·13, 1·14) |
| 50–64 | Mild Frailty | 3·10 (2·75–3·50) | 3·04 (2·69–3·44) | 1·58 (1·44–1·73) | 3·11 (2·95, 3·28) | 2·14 (2·05, 2·22) |
| | Moderate Frailty | 6·50 (5·60–7·55) | 6·28 (5·40–7·30) | 2·33 (2·04–2·66) | 5·10 (4·58, 5·66) | 3·74 (3·50, 4·00) |
| | Severe Frailty | 16·76 (13·86–20·26) | 16·41 (13·67–19·71) | 3·40 (2·75–4·21) | 5·67 (4·61, 6·98) | 6·61 (5·85, 7·48) |
| | eFI continuous | 1·13 (1·13–1·14) | 1·13 (1·12–1·14) | 1·06 (1·05–1·07) | 1·17 (1·16, 1·17) | 1·10 (1·09, 1·10) |
| 65–79 | Mild Frailty | 2·06 (1·90–2·24) | 2·26 (2·04–2·50) | 1·49 (1·37–1·61) | 2·49 (2·38, 2·61) | 1·63 (1·58, 1·69) |
| | Moderate Frailty | 4·55 (4·17–4·97) | 4·67 (4·20–5·20) | 1·98 (1·78–2·19) | 3·58 (3·34, 3·84) | 2·49 (2·39, 2·60) |
| | Severe Frailty | 10·33 (9·26–11·51) | 10·52 (9·33–11·85) | 2·80 (2·43–3·23) | 3·16 (2·85, 3·52) | 3·84 (3·59, 4·10) |
| | eFI continuous | 1·11 (1·10–1·11) | 1·11 (1·10–1·11) | 1·05 (1·04–1·05) | 1·12 (1·11, 1·12) | 1·07 (1·06, 1·07) |
| ≥80 | Mild Frailty | 2·02 (1·84–2·21) | 1·92 (1·68–2·20) | 1·52 (1·33–1·74) | 1·59 (1·49, 1·70) | 1·39 (1·32, 1·46) |
| | Moderate Frailty | 3·22 (2·94–3·54) | 3·36 (2·94–3·83) | 2·10 (1·84–2·40) | 2·23 (2·07, 2·41) | 1·81 (1·71, 1·92) |
| | Severe Frailty | 5·48 (4·96–6·05) | 5·89 (5·16–6·72) | 2·50 (2·16–2·90) | 2·26 (2·07, 2·46) | 2·28 (2·14, 2·43) |
| | eFI continuous | 1·08 (1·07–1·08) | 1·08 (1·07–1·08) | 1·04 (1·04–1·05) | 1·05 (1·05, 1·05) | 1·04 (1·04, 1·04) |

Estimates are presented as hazard ratios (HRs), odds ratios (ORs), and incidence rate ratios (IRRs) with 95% confidence intervals (CIs) for the electronic frailty index (eFI). HRs were derived from Cox proportional hazards models, and ORs and IRRs from hurdle negative binomial models. All models were adjusted for sex. For categorical eFI predictors, estimates are relative to the non-frail within each subgroup.

**Supplementary Table 10. Model fit statistics for severe infections and fractures in count models.**

| Model | Severe Infections (N=142229) | | Fractures (N = 142209) | |
|---|---|---|---|---|
| | AIC | BIC | AIC | BIC |
| **Base** | 74609 | 74679 | 105868 | 105937 |
| **eFI** | | | | |
| Categorical | 71448 | 71576 | 105100 | 105229 |
| Continuous | 71095 | 71184 | 104905 | 104994 |
| **HFRS** | | | | |
| Categorical | 73946 | 74054 | 105633 | 105742 |
| Continuous | 73586 | 73674 | 105389 | 105479 |
| **CCI** | | | | |
| Categorical | 72980 | 73108 | 105736 | 105865 |
| Continuous | 73316 | 73405 | 105785 | 105874 |

Akaike information criterion (AIC) and Bayesian information criterion (BIC) are presented for hurdle negative binomial models of frequencies of severe infections and fractures. The base model includes age and sex as predictors only. Additional models include the baseline electronic frailty index (eFI), Hospital Frailty Risk Score (HFRS), or Charlson Comorbidity Index (CCI) added to the base model to assess their relative model fit compared with the base model. Lower AIC and BIC values indicate better model fit.

**Supplementary Table 11. Model fit statistics for healthcare utilization.**

| Model | AIC | BIC |
|---|---|---|
| **Base** | 834202 | 834272 |
| **eFI** | | |
| Categorical | 817245 | 817376 |
| Continuous | 808816 | 808907 |
| **HFRS** | | |
| Categorical | 831629 | 831740 |
| Continuous | 827868 | 827959 |
| **CCI** | | |
| Categorical | 827214 | 827346 |
| Continuous | 828609 | 828700 |

Models were fitted using hurdle negative binomial regression. The base model included baseline age and sex, and additional models included eFI, HFRS, or CCI added separately to the base model to examine incremental model fit. Model performance was evaluated using the Akaike Information Criterion (AIC) and Bayesian Information Criterion (BIC), where lower values indicate better model fit.

CCI, Charlson comorbidity index; eFI, electronic frailty index; HFRS, hospital frailty risk score.

**Supplementary Table 12. Hold-out validation based on model concordance for models on all-cause mortality, severe infections, and fractures.**

| | | Training set | | | | Test set | | | |
|---|---|---|---|---|---|---|---|---|---|
| | | N | events | C-index | SE | N | events | C-index | SE |
| All-cause mortality | eFI | | | | | | | | |
| | Continuous | 87412 | 6295 | 0·8597 | 0·0037 | 37462 | 2628 | 0·8582 | 0·0057 |
| | Categorical | 87412 | 6295 | 0·8580 | 0·0037 | 37462 | 2628 | 0·8574 | 0·0057 |
| | HFRS | | | | | | | | |
| | Continuous | 87412 | 6295 | 0·8400 | 0·0037 | 37462 | 2628 | 0·8406 | 0·0057 |
| | Categorical | 87412 | 6295 | 0·8368 | 0·0037 | 37462 | 2628 | 0·8373 | 0·0057 |
| | CCI | | | | | | | | |
| | Categorical | 87412 | 6295 | 0·8498 | 0·0037 | 37462 | 2628 | 0·8483 | 0·0057 |
| | Continuous | 87412 | 6295 | 0·8479 | 0·0037 | 37462 | 2628 | 0·8474 | 0·0057 |
| Severe infections | eFI | | | | | | | | |
| | Continuous | 99561 | 4509 | 0·7655 | 0·0043 | 42668 | 1984 | 0·7554 | 0·0065 |
| | Categorical | 99561 | 4509 | 0·7564 | 0·0043 | 42668 | 1984 | 0·7434 | 0·0065 |
| | HFRS | | | | | | | | |
| | Continuous | 99561 | 4509 | 0·7187 | 0·0043 | 42668 | 1984 | 0·7101 | 0·0065 |
| | Categorical | 99561 | 4509 | 0·7070 | 0·0043 | 42668 | 1984 | 0·6981 | 0·0065 |
| | CCI | | | | | | | | |
| | Categorical | 99561 | 4509 | 0·7312 | 0·0043 | 42668 | 1984 | 0·7264 | 0·0065 |
| | Continuous | 99561 | 4509 | 0·7243 | 0·0043 | 42668 | 1984 | 0·7212 | 0·0065 |
| Fractures | eFI | | | | | | | | |
| | Continuous | 99547 | 6861 | 0·6171 | 0·0035 | 42662 | 3014 | 0·6177 | 0·0053 |
| | Categorical | 99547 | 6861 | 0·6057 | 0·0035 | 42662 | 3014 | 0·6039 | 0·0053 |
| | HFRS | | | | | | | | |
| | Continuous | 99547 | 6861 | 0·6005 | 0·0035 | 42662 | 3014 | 0·5989 | 0·0053 |
| | Categorical | 99547 | 6861 | 0·5862 | 0·0035 | 42662 | 3014 | 0·5847 | 0·0053 |
| | CCI | | | | | | | | |
| | Categorical | 99547 | 6861 | 0·5850 | 0·0035 | 42662 | 3014 | 0·5819 | 0·0053 |
| | Continuous | 99547 | 6861 | 0·5833 | 0·0035 | 42662 | 3014 | 0·5801 | 0·0053 |

Cox proportional hazards models were fitted for each outcome with adjustment for age and sex. Models included the electronic frailty index (eFI), Hospital Frailty Risk Score (HFRS), and Charlson Comorbidity Index (CCI), modelled as continuous and categorical predictors. Model discrimination was assessed using the concordance index (C-index) with its standard error (SE). Performance was evaluated using a 70% training and 30% test data split. Train values represent model discrimination estimated in the training dataset, and test values represent predictive performance in the held-out test dataset. The number of individuals (N) and number of events for each outcome are shown for both datasets.

**Supplementary Table 13. Cross-validated performance for severe infections and fractures based on count models.**

| Model | Severe Infections | | | | Fractures | | | |
|---|---|---|---|---|---|---|---|---|
| | Train (N = 99561) | | Test (N = 42668) | | Train (N = 99547) | | Test (N = 42662) | |
| | RMSE | MAE | RMSE | MAE | RMSE | MAE | RMSE | MAE |
| **Base** | 1·0660 | 0·2675 | 1·0642 | 0·2661 | 1·3106 | 0·3806 | 1·2680 | 0·3771 |
| **eFI** | | | | | | | | |
| Categorical | 1·0549 | 0·2547 | 1·0513 | 0·2517 | 1·3078 | 0·3772 | 1·2654 | 0·3731 |
| Continuous | 1·0543 | 0·2535 | 1·0501 | 0·2509 | 1·3080 | 0·3768 | 1·2655 | 0·3724 |
| **CCI** | | | | | | | | |
| Categorical | 1·0615 | 0·2614 | 1·0593 | 0·2599 | 1·3102 | 0·3801 | 1·2680 | 0·3770 |
| Continuous | 1·0648 | 0·2629 | 1·0626 | 0·2620 | 1·3104 | 0·3803 | 1·2677 | 0·3768 |
| **HFRS** | | | | | | | | |
| Categorical | 1·0637 | 0·2642 | 1·0627 | 0·2629 | 1·3096 | 0·3793 | 1·2679 | 0·3758 |
| Continuous | 1·0647 | 0·2636 | 1·0627 | 0·2621 | 1·3096 | 0·3785 | 1·2685 | 0·3748 |

Models were fitted using hurdle negative binomial regression for severe infections and fractures counts. The base model included baseline age and sex, and additional models included eFI, HFRS, or CCI added separately to the base model. Predictive performance was assessed using root mean square error (RMSE) and mean absolute error (MAE) using a 70% training and 30% test data split. Train values represent 10-fold cross-validated performance on the training data, and test values represent predictive performance on the held-out test data.

**Supplementary Table 14. Cross-validated model performance for the total healthcare use models.**

| Model | Train (N = 124715) | | Test (N = 53448) | |
|---|---|---|---|---|
| | RMSE | MAE | RMSE | MAE |
| **Base** | 11·3790 | 6·0490 | 11·5146 | 6·0738 |
| **eFI** | | | | |
| Categorical | 10·8745 | 5·6098 | 11·0081 | 5·6261 |
| Continuous | 11·0612 | 5·5640 | 11·2107 | 5·5841 |
| **CCI** | | | | |
| Categorical | 11·1671 | 5·8985 | 11·3342 | 5·9352 |
| Continuous | 12·4594 | 6·1507 | 12·4077 | 6·1707 |
| **HFRS** | | | | |
| Categorical | 11·2931 | 5·9774 | 11·4468 | 6·0143 |
| Continuous | 11·9603 | 6·0333 | 12·1227 | 6·0786 |

Models were fitted using hurdle negative binomial regression. The base model included age and sex, and additional models included eFI, HFRS, or CCI added individually to the base model. Predictive performance was assessed using root mean square error (RMSE) and mean absolute error (MAE) using a 70% training and 30% test data split. Train values represent 10-fold cross-validated performance on the training data, and test values represent predictive performance on the held-out test data.
eFI, electronic frailty index; HFRS, hospital frailty risk score; CCI, Charlson comorbidity index.

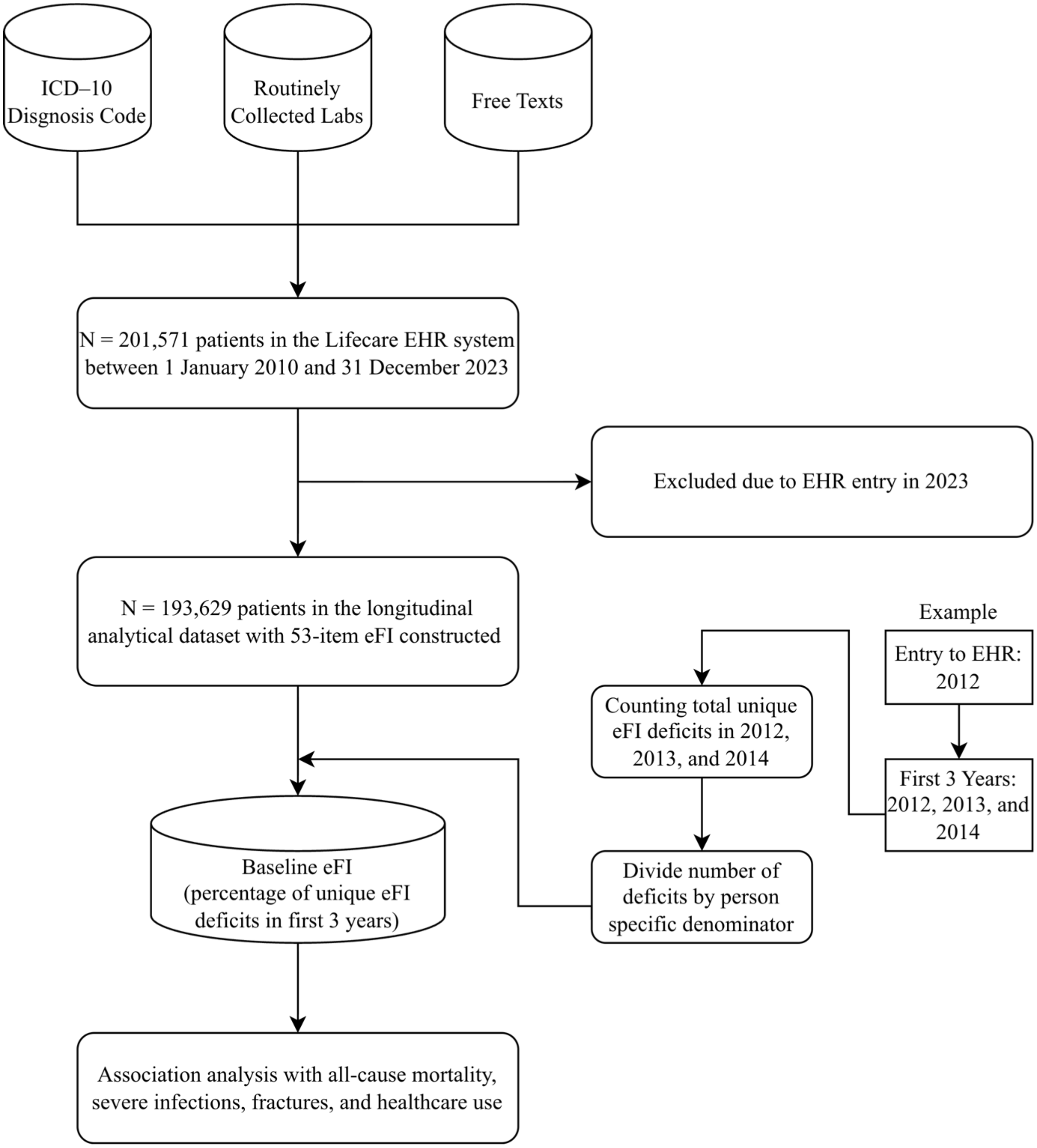


**Supplementary Figure 1. Flowchart of baseline eFI construction.**
eFI, electronic frailty index; EHR, electronic health record; ICD-10, International Classification of Diseases, 10th Revision; N, number of individuals.

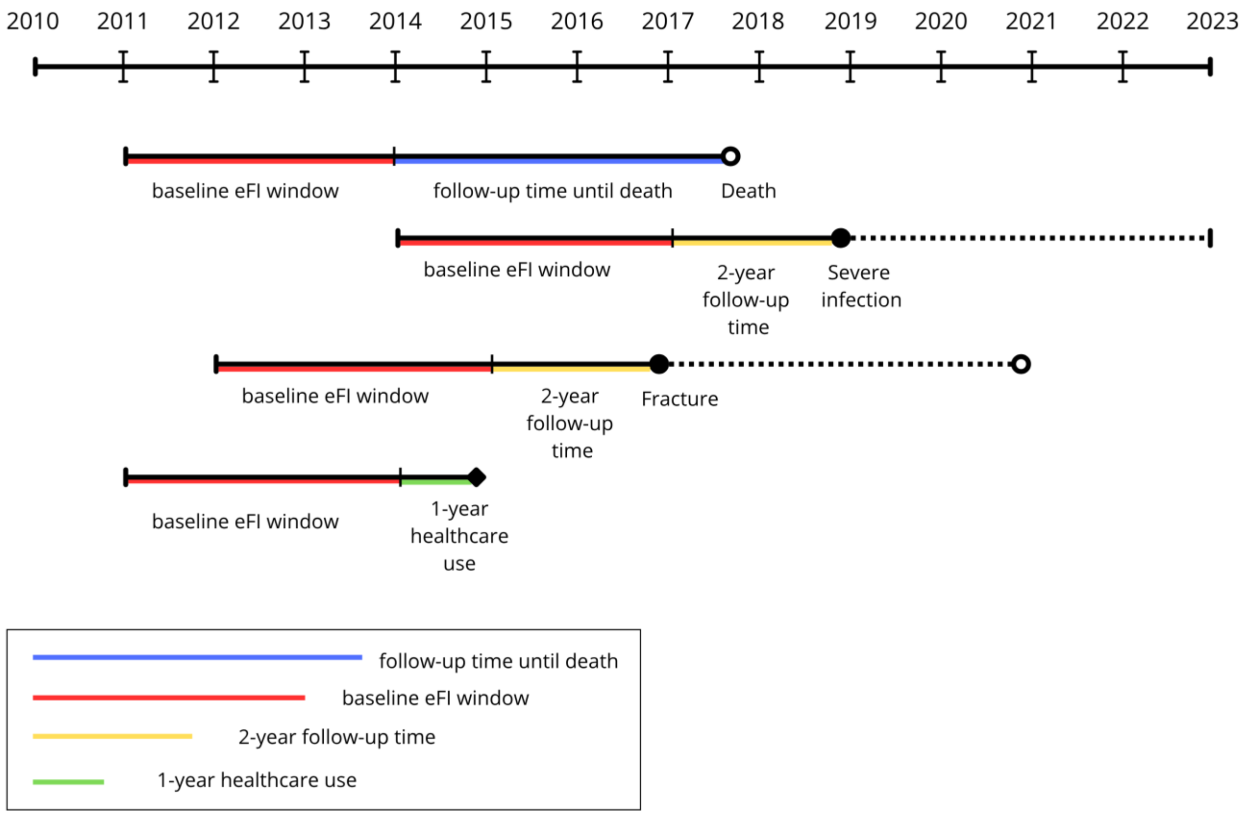


**Supplementary Figure 2. Study design and time windows for baseline eFI assessment and outcome follow-up.** The timeline illustrates the definition of baseline eFI and follow-up periods within the observation period (2010–2023).

The baseline eFI window (red) was used to compute eFI prior to outcome assessment. For mortality, follow-up extended from the end of baseline until death or censoring (blue).

For the other clinical outcomes (severe infections and fractures), a fixed 2-year follow-up period was applied after baseline (yellow), with individuals censored thereafter (dotted lines). For healthcare utilization, outcomes were assessed within 1 year after baseline (green).

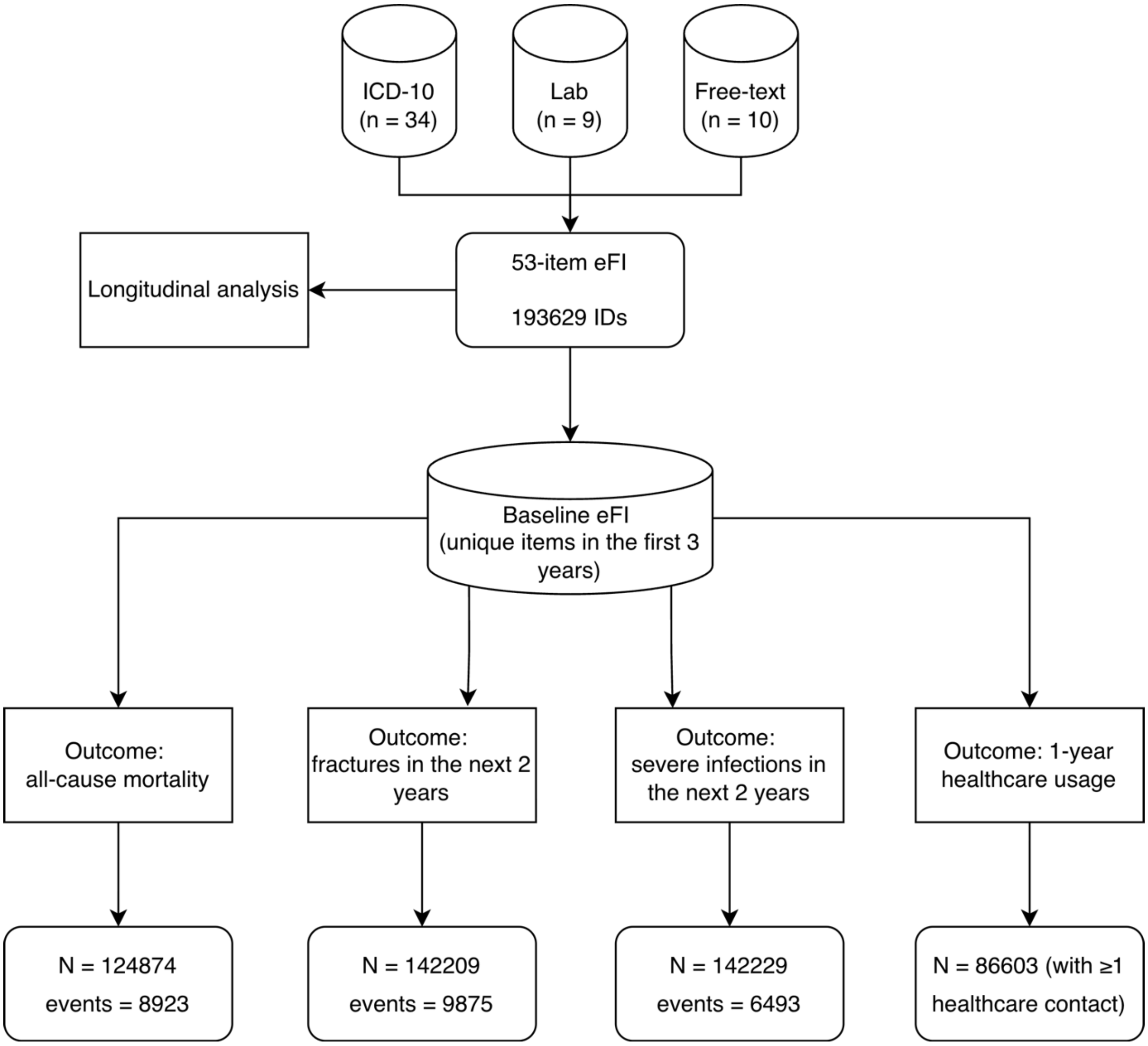


**Supplementary Figure 3. Outcome-specific analytical samples derived from the baseline eFI cohort.**
Starting from the cohort with available baseline eFI, outcome-specific analytical samples were defined according to eligibility and follow-up requirements. Sample sizes and number of events are shown for each outcome: mortality, fractures within 2 years, severe infections within 2 years, and 1-year healthcare use.
eFI, electronic frailty index; EHR, electronic health record; ICD-10, International Classification of Diseases, 10th revision; ID, participant identifier; n, number of items, N, number of individuals.

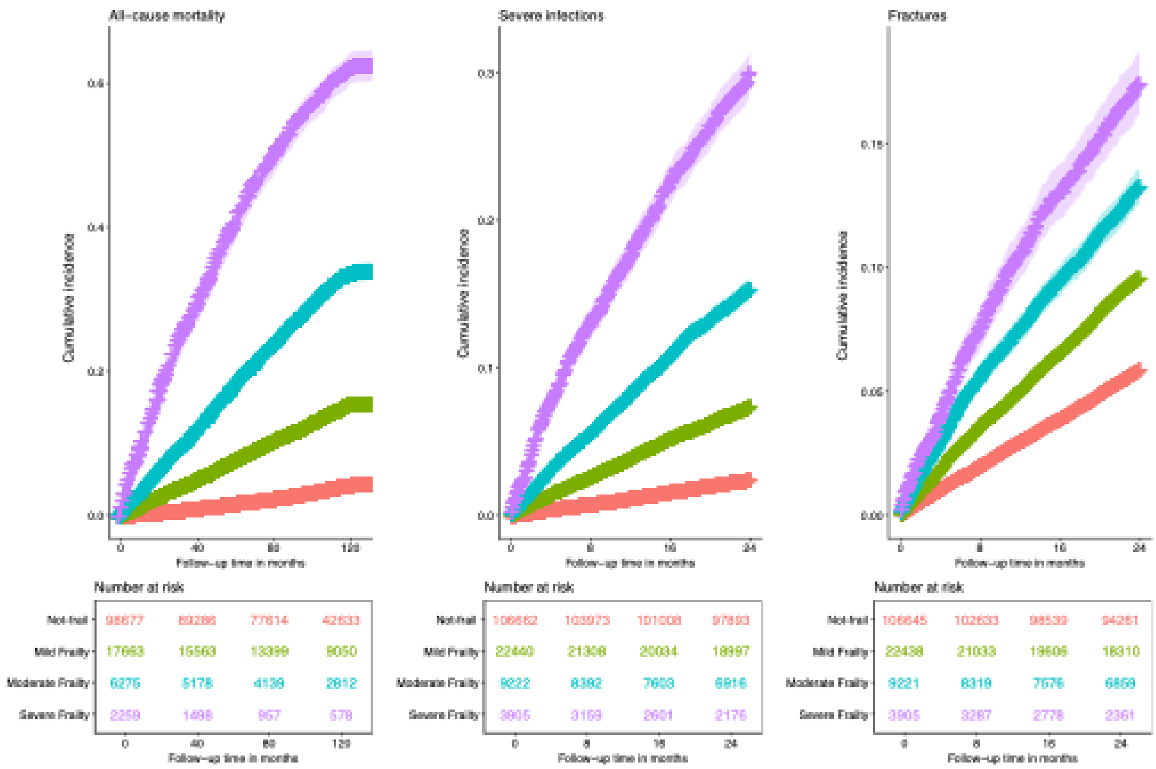


**Supplementary Figure 4. Kaplan–Meier curves for all-cause mortality, severe infections, and fractures by eFI categories.**

Individuals at risk are shown below each panel. The left panel shows all-cause mortality over 120 months of follow-up, whereas severe infections and fractures are shown over 24 months.
